# Robust clinical detection of SARS-CoV-2 variants by RT-PCR/MALDI-TOF multi-target approach

**DOI:** 10.1101/2021.09.09.21263348

**Authors:** Matthew M. Hernandez, Radhika Banu, Ana S. Gonzalez-Reiche, Adriana van de Guchte, Zenab Khan, Paras Shrestha, Liyong Cao, Feng Chen, Huanzhi Shi, Ayman Hanna, Hala Alshammary, Shelcie Fabre, Angela Amoako, Ajay Obla, Bremy Alburquerque, Luz Helena Patiño, Juan David Ramírez, Robert Sebra, Melissa R. Gitman, Michael D. Nowak, Carlos Cordon-Cardo, Ted E. Schutzbank, Viviana Simon, Harm van Bakel, Emilia Mia Sordillo, Alberto E. Paniz-Mondolfi

**Affiliations:** Department of Pathology, Molecular, and Cell-Based Medicine, Icahn School of Medicine at Mount Sinai, New York, NY 10029, USA; Department of Genetics and Genomic Sciences, Icahn School of Medicine at Mount Sinai, New York, NY 10029, USA; Department of Microbiology, Icahn School of Medicine at Mount Sinai, New York, NY 10029, USA; The Graduate School of Biomedical Sciences, Icahn School of Medicine at Mount Sinai, New York, NY 10029, USA; Centro de Investigaciones en Microbiología y Biotecnología-UR (CIMBIUR), Facultad de Ciencias Naturales, Universidad del Rosario, Bogotá, Colombia; Icahn Institute for Data Science and Genomic Technology, Icahn School of Medicine at Mount Sinai, New York, NY 10029, USA; Black Family Stem Cell Institute, Icahn School of Medicine at Mount Sinai, New York, NY 10029, USA; Sema4, a Mount Sinai venture, Stamford, CT 06902, USA; Senior Scientific Affairs Manager, Infectious Diseases, Agena Bioscience, San Diego, CA 92121, USA; Division of Infectious Diseases, Department of Medicine, Icahn School of Medicine at Mount Sinai, New York, NY 10029, USA; The Global Health and Emerging Pathogens Institute, Icahn School of Medicine at Mount Sinai, New York, NY 10029, USA

**Keywords:** RT-PCR, MALDI-TOF, SARS-CoV-2, B.1.1.7, variants, diagnostic, dropout

## Abstract

The COVID-19 pandemic sparked rapid development of SARS-CoV-2 diagnostics. However, emerging variants pose the risk for target dropout and false-negative results secondary to primer/probe binding site (PBS) mismatches. The Agena MassARRAY^®^ SARS-CoV-2 Panel combines RT-PCR and MALDI-TOF mass-spectrometry to probe for five targets across *N* and *ORF1ab* genes, which provides a robust platform to accommodate PBS mismatches in divergent viruses. Herein, we utilize a deidentified dataset of 1,262 SARS-CoV-2-positive specimens from Mount Sinai Health System (New York City) from December 2020 through April 2021 to evaluate target results and corresponding sequencing data. Overall, the level of PBS mismatches was greater in specimens with target dropout. Of specimens with N3 target dropout, 57% harbored an A28095T substitution that is highly-specific for the alpha (B.1.1.7) variant of concern. These data highlight the benefit of redundancy in target design and the potential for target performance to illuminate the dynamics of circulating SARS-CoV-2 variants.

## Introduction

Molecular diagnostic assays for severe acute respiratory syndrome coronavirus-2 (SARS-CoV-2), the etiologic agent of coronavirus disease 2019 (COVID-19), utilize nucleic acid amplification test (NAAT) methods to assess for presence of viral nucleic acids in clinical specimens. These assays rely on primers and probes targeting one or more viral gene regions including open reading frame 1ab (*ORF1ab*), open reading frame 8 (*ORF 8)*, nucleocapsid (*N*), spike (*S*), and envelope (*E*) ^1^. These targets have been designed primarily based on sequences from virus strains that circulated early in the pandemic, including the reference genome collected from Wuhan, China in January, 2020 ^2–4^.

In addition to the quantity of viral nucleic acids in a clinical specimen, the diagnostic and analytic capabilities of NAATs depend on the complementarity of primers and probes to viral genome sequences to reliably amplify targets of interest. As a result, binding of primers and probes can be impacted by progressive accumulation of changes in the viral genomes at primer binding sites (PBSs). Indeed, mismatches in PBSs – particularly the 2-3 nucleotides at the 3’ end of the oligonucleotide – can result in reduced binding and subsequent failure to amplify (termed “dropout” in diagnostic NAAT assays) ^5–8^. In fact, SARS-CoV-2 has diversified over the past 18 months, and mutations in the *N, S*, and *E* genes have been reported in viruses from specimens with corresponding target dropout during testing on commercial NAAT-based diagnostic platforms ^9–15^. Moreover, *in silico* analyses have utilized publicly-available SARS-CoV-2 genome sequences to identify mutations in circulating viral variants that have the potential to interfere with diagnostic targets ^1,5,16–18^. These findings highlight the potential diagnostic challenge as increasingly diverse SARS-CoV-2 lineages (e.g., B.1.1.7) continue to emerge globally ^19,20^.

To limit the risk of false-negative results, most NAAT assays for SARS-CoV-2 that currently have emergency use authorization (EUA) from the US Food and Drug Administration (FDA) utilize two or more diagnostic targets ^21,22^. We recently reported the analytic performance of the Agena MassARRAY^®^ SARS-CoV-2 Panel which combines RT-PCR and matrix-assisted laser desorption/ionization time-of-flight (MALDI-TOF) technologies to detect SARS-CoV-2 ^23^. The Agena MassARRAY^®^ platform probes for five distinct targets in the *ORF1ab* and *N* viral genes ^24^, providing a robust platform for diagnosis of SARS-CoV-2 in clinical specimens despite the emergence of virus strains that have accumulated mutations that can interfere with some diagnostic targets. We evaluated the pattern of target detections for SARS-CoV-2-positive specimens collected at the Mount Sinai Health System (MSHS) to interrogate the impact of viral genetic variation on this diagnostic platform.

To do this, we compared detection of Agena diagnostic targets and genomic sequence data for SARS-CoV-2-positive specimens that were deidentified and banked as part of our Pathogen Surveillance Program (MSHS PSP) at the Icahn School of Medicine at Mount Sinai (ISMMS), which has been previously described ^25^. Complete viral genomes underwent phylogenetic analyses to characterize emergent evolutionary lineages among the SARS-CoV-2-positive specimens at MSHS (manuscript in preparation). For this analysis, we utilized a dataset comprised of 1,262 viral genomes recovered from deidentified clinical specimens collected from patients seeking care at the Mount Sinai Health System from December 1, 2020 through April 24, 2021. We identified PBS mismatches associated with lineage-specific substitutions in SARS-CoV-2 variants of concern (VOC) that resulted in Agena MassARRAY^®^ platform target dropout.

## Materials and Methods

### Ethics statement

This study was reviewed and approved by the Institutional Review Board of the Icahn School of Medicine at Mount Sinai (HS#13-00981).

### SARS-CoV-2 specimen collection and testing

Upper respiratory tract (e.g., nasopharyngeal, anterior nares) and saliva specimens collected for SARS-CoV-2 testing underwent diagnostic testing in the MSHS Clinical Microbiology Laboratory (CML), which is certified under Clinical Laboratory Improvement Amendments of 1988 (CLIA), 42 U.S.C. §263a and meets requirements to perform high-complexity tests. For this study, we retrospectively utilized deidentified data available for diagnostic specimens tested on the Agena MassARRAY^®^ SARS-CoV-2 Panel and MassARRAY^®^ System (Agena, CPM384) platform during the study period.

As previously described, prior to SARS-CoV-2 testing, saliva specimens underwent an initial processing step involving a 15 minute incubation at 55°C prior to RNA extraction ^23^. Upper respiratory specimens did not undergo any pre-processing prior to testing. RNA was extracted from 300μL of each specimen using the chemagic™ Viral DNA/RNA 300 Kit H96 (PerkinElmer, CMG-1033-S) on the automated chemagic™ 360 instrument (PerkinElmer, 2024-0020) per the manufacturer’s protocol. The MS2 phage RNA internal control (IC) was included in all extraction steps. The extracted RNA underwent RT-PCR with iPLEX^®^ Pro chemistry to amplify different Agena targets, per the manufacturer’s protocol. After inactivation of unincorporated dNTPs by treatment with shrimp alkaline phosphatase (SAP), a sequence-specific primer extension step was performed, in which a mass-modified terminator nucleotide was added to the probe, using supplied extension primers and iPLEX^®^ Pro reagents.

Extension products (analytes) were desalted, transferred to a SpectroCHIP^®^ Array (silicon chip with pre-spotted matrix crystal) and loaded into the MassARRAY^®^ Analyzer (a MALDI-TOF mass spectrometer). The analyte/matrix co-crystals were irradiated by a laser inducing desorption and ionization, and positively charged molecules accelerated into a flight tube towards a detector. Separation occurred by time-of-flight, which is proportional to molecular mass. After data processing, a spectral fingerprint was generated for each analyte that characterizes the mass/charge ratio and relative intensity of the molecules. Data acquired by the MassARRAY^®^ Analyzer was processed with the MassARRAY^®^ Typer software and SARS-CoV-2 Report software. The assay detects five viral targets: three in the nucleocapsid (*N*) gene (N1, N2, N3) and two in the *ORF1ab* gene (ORF1A, ORF1AB). If the IC was detected, results were interpreted as positive if ≥ 2 targets were detected or negative if < 2 targets were detected. If no IC and no targets were detected, the result was invalid and required repeat testing of the specimen before reporting. If IC was detected and no targets were detected, the sample was interpreted as negative.

Overall, 86,781 upper respiratory and saliva specimens underwent clinical testing in the CML at MSHS, during the period from December 1, 2020 through April 24, 2021. Of those specimens, 2,062 tested positive for SARS-CoV-2. A subset of 1,262 specimens were deidentified, related data was entered in to the MSHS PSP database, and underwent SARS-CoV-2 next-generation sequencing as previously described ^25,26^.

### SARS-CoV-2 sequencing, assembly and phylogenetic analyses

SARS-CoV-2 viral RNA underwent reverse transcription, PCR amplification and next-generation sequencing followed by genome assembly and lineage assignment using a phylogenetic-based nomenclature as described by Rambaut et al. ^27^ using the PANGOLIN tool, version 2021-04-28 ^28^ as previously described ^25,26^. Ultimately, this yielded 1,176 complete genomes (≥95% completeness) and 86 partial genomes (<95% completeness).

### Agena target sequence alignment

Agena MassARRAY^®^ target detection results were matched to the corresponding genome sequences. Primer and probe sequences for each Agena target were obtained from published FDA EUA documentation for the Agena MassARRAY^®^ SARS-CoV-2 Panel (**Supplemental Table S1**) ^24^. We generated reverse-complement sequences for reverse primers for all five targets and probes that are designed in the reverse orientation (e.g., N1-N3). An unaligned FASTA file including sequence data for the clinical specimens and the Wuhan-Hu-1 reference sequence (NCBI nucleotide: NC_045512.2 (Genbank: MN908947.3)) was generated for each of the fifteen primers/probes. The Multiple Alignment using Fast Fourier Transform (MAFFT) platform ^29,30^ which is publicly available for use online (https://mafft.cbrc.jp/alignment/server/add_fragments.html?frommanualnov6) was used to align each file. To enable inclusion of incomplete genomes that had intact regions sequenced at PBSs, we did not remove uninformative sequences (e.g., with ambiguous letters). Otherwise, the default settings were used to align all sequences to the reference genome, which generated a resulting FASTA alignment file for each primer and probe sequence.

### Sequence variation in primer/probe target regions

To identify mismatches in the primer and probe regions of the viral genomes, FASTA alignment files were processed locally in a Bash environment. Custom Unix-code (https://github.com/AceM1188/SACOV_primer-probe_analyses) was used to identify mismatches at each nucleotide position within each primer and probe sequence ^31^. A tab-delimited output file that identified mismatches by primer/probe nucleotide position across the viral genome sequences was generated for each alignment.

Note that for the viral genome sequences with stretches of Ns that corresponded with the PBS, mismatches could not be called, and these sequences were excluded from the mismatch counting for the given primer/probe. In addition, genomes with gaps that spanned the entire region of a PBS were excluded from the analyses for the given primer/probe.

Mismatches by position in PBS regions of forward/reverse primer and probe sequences were manually counted on Microsoft Excel v16.48. To account for differences in completeness of consensus genomes, the number of PBS mismatches was normalized to the number of nucleotides in the PBS of each specimen consensus sequence.

### Statistical analyses

For statistical comparison of fraction of PBS with mismatches in genomes with detected targets versus those with dropout targets, normality was assessed by D’Agostino and Pearson test (GraphPad Prism 9.1.0), which indicated that all distributions were non-parametric; thus, a Mann-Whitney test (two-tailed) was performed (GraphPad). To determine if specific mismatches were associated with specific target dropout results, specimens were grouped by (1) presence or absence of the mismatch of interest (in the setting of no other mismatches) and (2) detection or dropout of the target of interest – which resulted in a 2×2 contingency table which underwent association testing by Fisher’s exact test.

### Display Items

All figures are original and were generated using the GraphPad Prism software 9.1.0, R software package ggplot2, NCBI Multiple Sequence Alignment Viewer v.1.17.0 (https://www.ncbi.nlm.nih.gov/tools/msaviewer/), and finished in Adobe Illustrator 2021 (v.25.2.1).

## Results

Overall, of the 2,062 SARS-CoV-2-positive specimens, 1,274 (62%) had all five targets detected with the remaining having one (n = 419) or more (n = 369) targets dropout. For the subset of 1,262 SARS-CoV-2-positive specimens sequenced in our study, all five diagnostic targets were detected in 943 (75%), with the remaining having one (n = 227) or more (n = 92) target dropout (**Supplemental Table S2**). When we calculated the target detection rate among these SARS-CoV-2-positive specimens by week, the ORF1AB target had the lowest average detection rate per week (0.87) followed by the N3 target (0.88) and the N2 target (0.94) (**Figure 1**). Notably, the N3 detection rate declined over time with the lowest detection occurring during the last four weeks of the timeframe studied (week ending April 3 (0.75) – April 24, 2021 (0.79)). Given these observations, we used the diagnostic data and corresponding genome sequences to identify mismatches to each primer/probe utilized by the Agena MassARRAY^®^ platform to determine the impact on target detection results.

**Figure 1.**
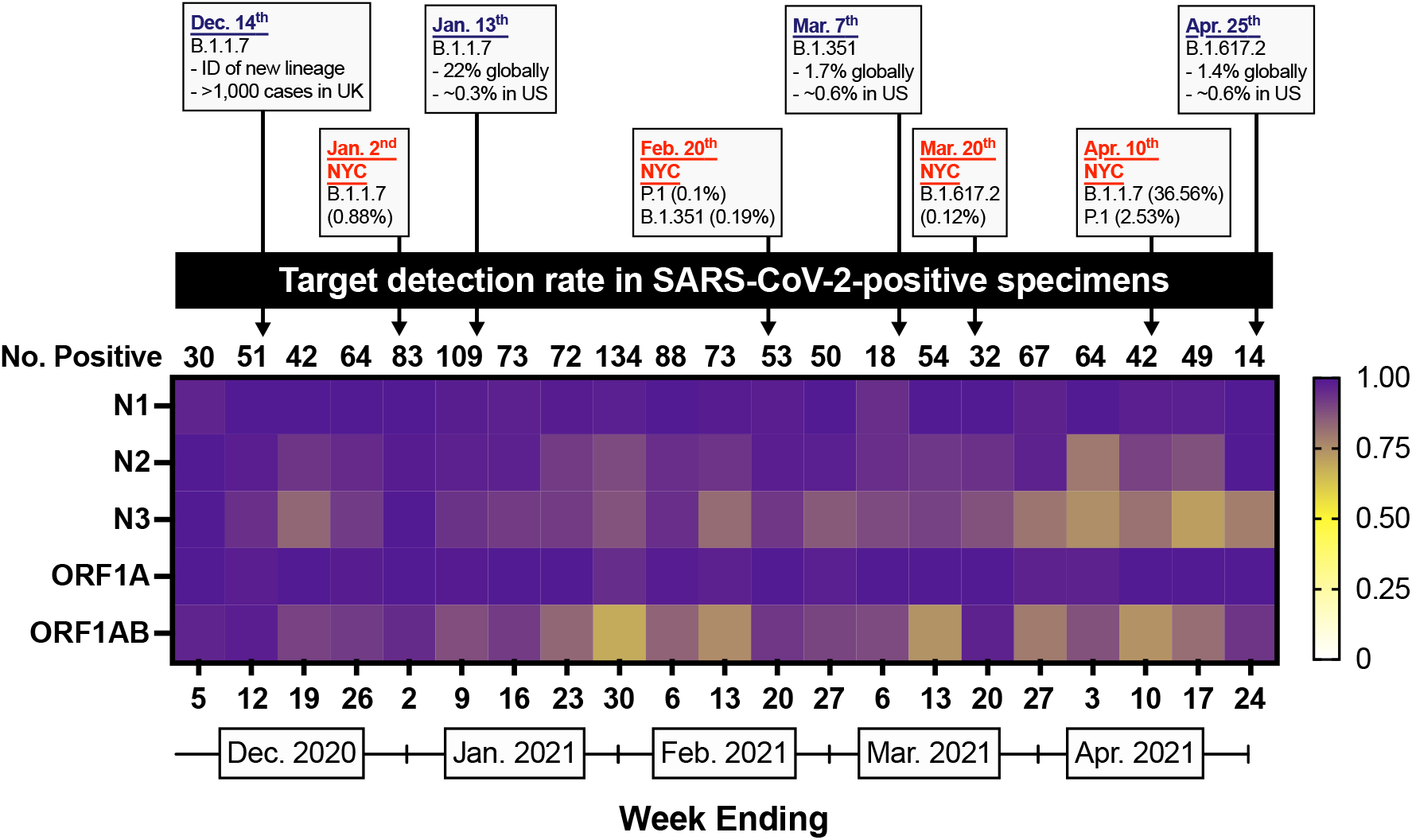
Agena target detection rate in SARS-CoV-2-positive specimens by week. Heatmap depicting the proportion of sequenced SARS-CoV-2-positive specimens that have detectable N1, N2, N3, ORF1A, or ORF1AB targets by week from December 1, 2020 through April 24, 2021. International, national, and global statistics are indicated by dates in purple font. NYC statistics are indicated by dates in red font. Data for epidemiologic events obtained from ^38,39,41,44,46^. The number of sequenced SARS-CoV-2-positive specimens per week is indicated above each week (column).

### Nucleotide mismatches across diagnostic targets

We aligned each forward primer, reverse primer, and probe sequence of the Agena MassARRAY^®^ SARS-CoV-2 Panel to the set of 1,262 SARS-CoV-2 genome sequences.

To examine the impact of mismatches on target results, we measured the number of mismatches (normalized to the number of nucleotides in the PBS; see methods) in specimens with detected and undetected target results (**Figure 2**). Detection of each of four targets (N1, N3, ORF1A, ORF1AB) was associated with perfect complementarity (0 mismatches) between the genome sequence and the respective target PBSs. Specifically, > 96% of specimens with either detectable N1 or N3 targets had perfect complementarity to the respective forward/reverse/probe PBS, and > 95% of specimens with detectable ORF1A or ORF1AB targets had perfect complementarity to the respective forward/reverse/probe PBS. The remaining specimens had – at most – only one mismatch to each of the target PBSs. The exception to this was the N2 target, for which, more specimens with detectable N2 target had mismatches to N2 forward (43%) and N2 reverse (39%) PBSs (**Figure 2B**). Indeed, up to four mismatches to the N2 forward and up to two mismatches to the N2 reverse PBSs were found in the specimens for which the N2 target was detected.

**Figure 2.**
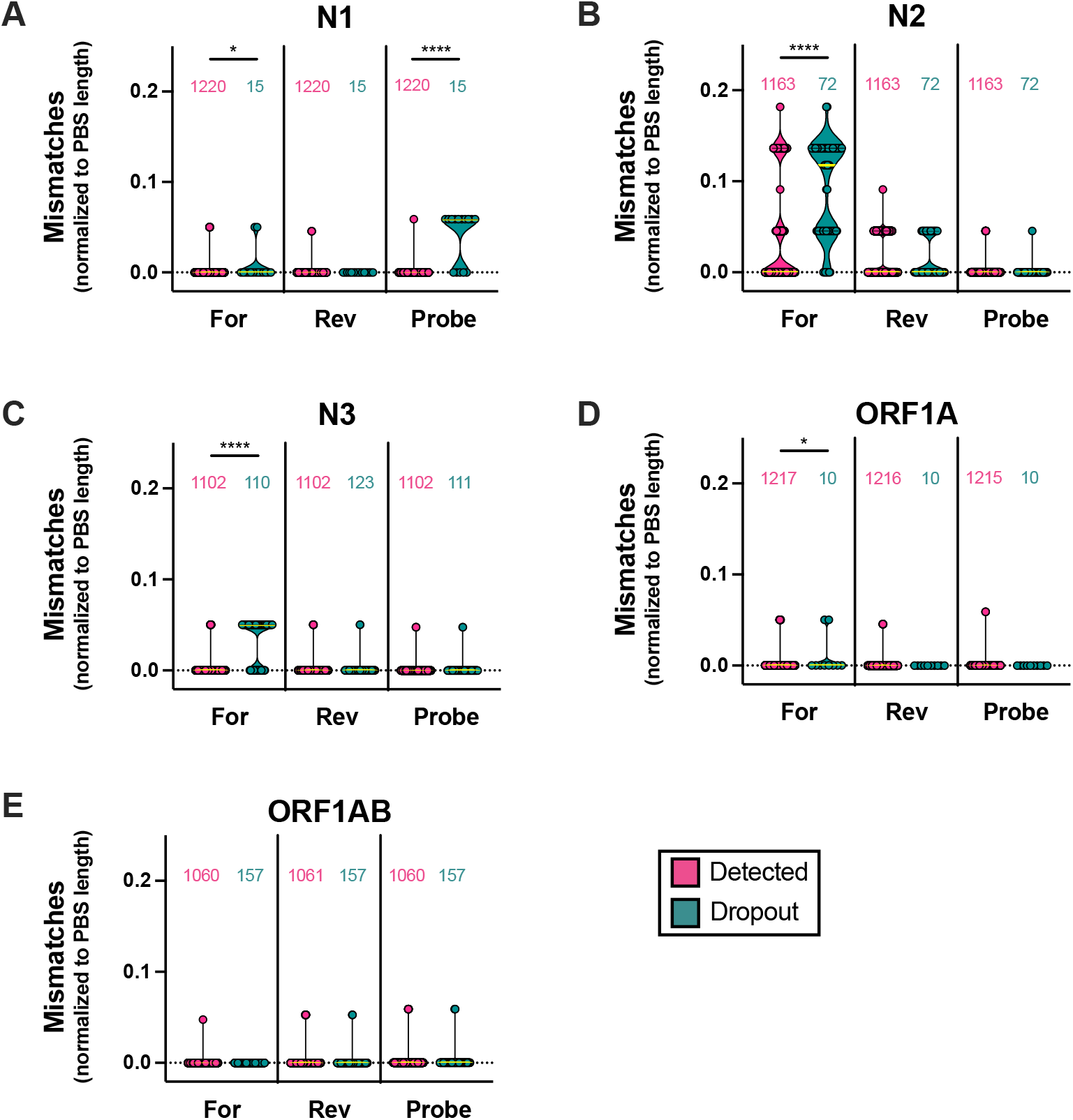
Impact of SARS-CoV-2 primer/probe binding site mismatches on Agena target detection results. Number of mismatches normalized to the number of nucleotides in primer/probe binding sites (PBS length) across five Agena MassARRAY^®^ diagnostic targets: **A:** N1, **B:** N2, **C:** N3, **D:** ORF1A, **E:** ORF1AB. Each point represents the calculated mismatches per specimen consensus genome for each target PBS. Violin plots represent the distribution as density of the points grouped by primer/probe sequence (forward (For), reverse (Rev), Probe) and by target detection result (detected (magenta), dropout (turquoise)). The number of genome sequences analyzed for mismatches are depicted above each violin plot. Medians are depicted as yellow lines. Bars above distributions reflect statistical comparison of underlying distributions by Mann-Whitney test. Asterisks reflect p-values (*, p < 0.05; ****, p < 0.0001).

When compared across target result groups, the number of mismatches was significantly higher in specimens with N1, N2, N3 and ORF1A target dropout (**Figure 2A-D**). In addition, we found the fraction of N1 probe PBS with mismatches was significantly higher in specimens with N1 target dropout than in those with detectable N1 (**Figure 2A**).

Because the position of mismatches within PBSs affect primer binding capabilities ^1,6–8^, we characterized the mismatch frequency by position of each primer/probe. Specifically, we measured the proportion of specimen genomes with a mismatch at each independent position along the full length of each target’s primer/probe (**Figure 3, Supplemental Figure S1**). From 5’ to 3’ direction, we found that among 15 specimens with N1 target dropout, 10 harbored single mismatches to the 4^th^ – 14^th^ basepair (bp) (SARS-CoV-2 genome positions 28714 – 28704) of the 17-bp-long N1 probe PBS (**Figure 3A**). Specifically, these mismatches reflected the following substitutions: G28714A (n = 1 specimens), G28713A (n = 2), C28709T (n = 2), C28706T (n = 1), G28704C (n = 3), G28704T (n = 1).

**Figure 3.**
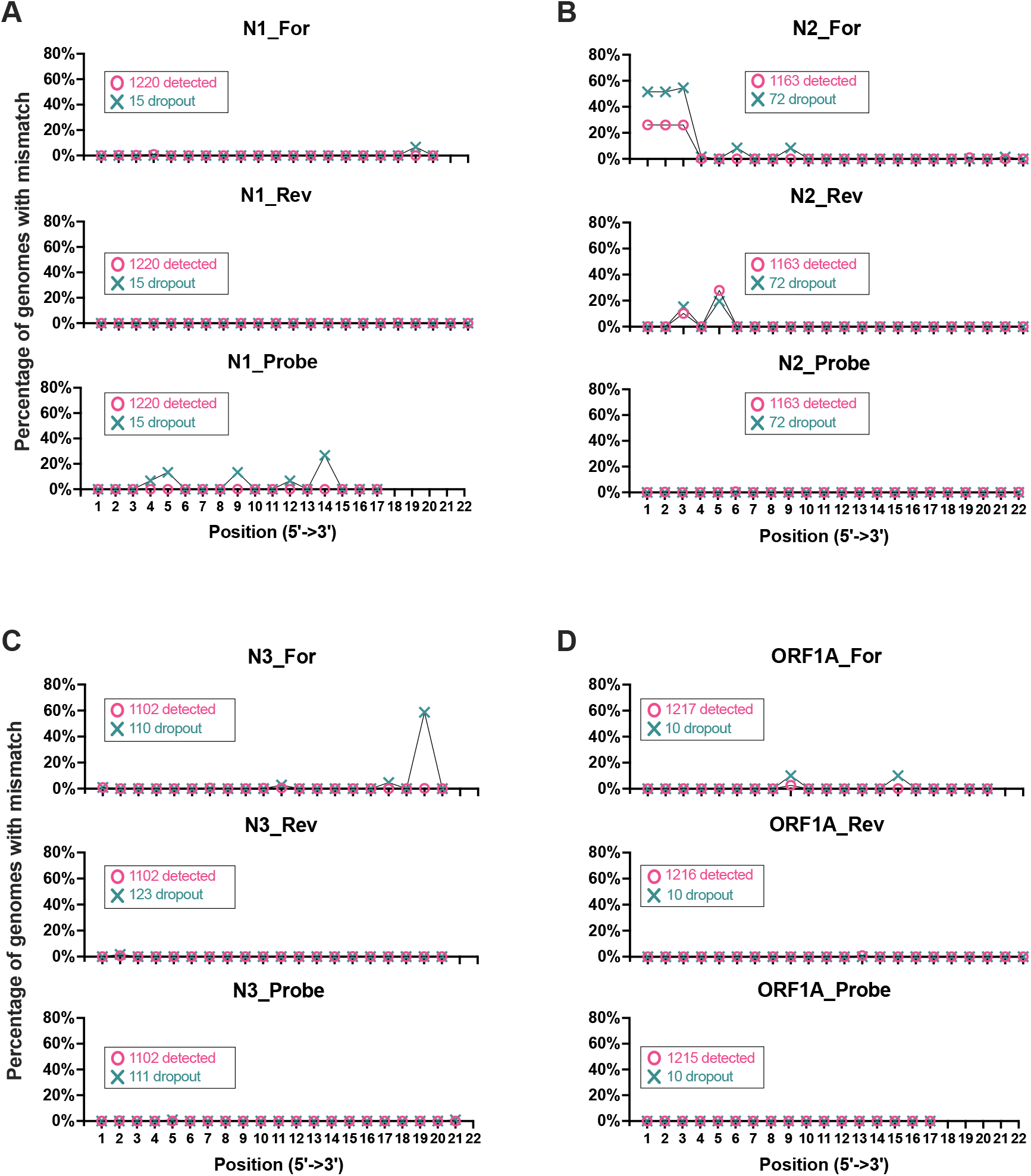
SARS-CoV-2 positional mismatches at target primer/probe binding sites. Line graphs depict the percentage of specimen genomes with mismatches at individual basepair positions across Agena MassARRAY^®^ target PBSs: **A:** N1, **B:** N2, **C:** N3, **D:** ORF1A. There are three plots for each target that correspond with the forward (For), reverse (Rev), and Probe binding sites. Two line plots are depicted for each binding site to depict mismatches in genomes from specimens that yielded a detected target result (magenta) or target dropout (turquoise). The percentage represents the number of genomes with mismatches at each position relative to the number of genome sequences detected or not detected by each target (annotated in each graph).

By contrast, mismatches in the 5’ end of the 22-bp-long N2 primer PBSs (forward, 1^st^ – 3^rd^ bp (28881 – 28883); reverse, 3^rd^ bp (28977) and 5^th^ bp (28975)) were identified in sequences that yielded both N2 target detection and dropout (**Figure 3B**). In 340 specimens with any one mismatch to the first 3 bp of the N2 forward primer, 336 (99%) harbored the concurrent substitutions G28881A, G28882A, and G28883C in the *N* gene. Of the 72 specimen genomes with N2 target dropout, 34 (47%) had this substitution trio. Although, this polymorphism was found in 304 (26%) of the 1,163 specimen genomes with N2 target detection, statistically, this represents a significant association of the GGG-to-AAC substitution with N2 target dropout (Fisher’s exact, p = 0.0002).

In addition, specimens that harbor mismatches to the 5’ end of the N2 reverse primer are the result of the C28977T or G28975A substitutions. However, of 466 specimens that harbor either substitution, only 1 had both suggesting these substitutions occur independently of one another. When grouped by N2 target detection result, neither substitution was significantly associated with N2 target dropout (Fisher’s exact, p ≥ 0.1351).

Interestingly, we found that of the 110 specimen genomes with N3 target dropout, 63 (57%) had a mismatch at the penultimate nucleotide towards the 3’ end in the 20-bp-long N3 forward primer (**Figure 3C**). All mismatches at this position are the result of a specific adenine-to-thymine substitution in *ORF8* (A28095T) of the SARS-CoV-2 genome. Of the 1,102 genomes with detected N3 target, only two harbored this mismatch; overall, this represents a statistically significant association of this positional mismatch with N3 target dropout (Fisher’s exact, p < 0.0001).

We also assessed whether the association of these mismatches with target dropout is maintained when the quantity of virus in the specimen is controlled. Although the Agena platform yields a qualitative diagnostic result, we have demonstrated previously that the number of detected targets is proportional to the quantity of virus in a given specimen ^23^. When we limit our dataset only to specimens for which all other (e.g., non-N3) targets are detected, the association of the A28095T substitution with N3 target dropout remains statistically significant (Fisher’s exact, p < 0.0001), indicating that N3 target dropout due to the A28095T substitution is independent of differences in virus concentration.

### Lineage-specific variation and target dropout

In order to assess whether target dropout was due to lineage-specific variation, we examined the phylogenetic lineages of genomes harboring distinct substitutions in our dataset. Among the 34 specimens with the concurrent GGG-to-AAC tri-nucleotide substitution and N2 target dropout, the earliest was from December 29, 2020 (PV24926) which belonged to the B.1.1.434 lineage. This polymorphism did not demonstrate bias to any one lineage in specimens that yielded N2 target dropout as it was found in specimens that mapped to 15 different lineages including B.1.1.7 (alpha, n = 11), B.1.1.434 (n = 6), and B.1.1 (n = 4) lineages.

We next examined the phylogenetic lineage of genomes harboring the A28095T substitution in our dataset to assess whether N3 target dropout was due to lineage-specific variation. We found that the earliest specimen with this substitution was from January 8, 2021 (specimen PV25263) and belonged to the B.1.1.7 lineage. Indeed, the substitution appeared in a subset of genomes of the B.1.1.7 lineage. Interestingly, of the 127 B.1.1.7 genomes, approximately half (n = 65) harbored the A28095T substitution while the remaining maintained the adenine at the position (**Figure 4**). Ninety-seven percent (63/65) of the B.1.1.7 specimens with the A20895T substitution demonstrated N3 target dropout. Furthermore, the converse was also true as almost all B.1.1.7 specimens with N3 target dropout (63/64 (98%)) had the A28095T substitution (**Figure 4A**).

**Figure 4.**
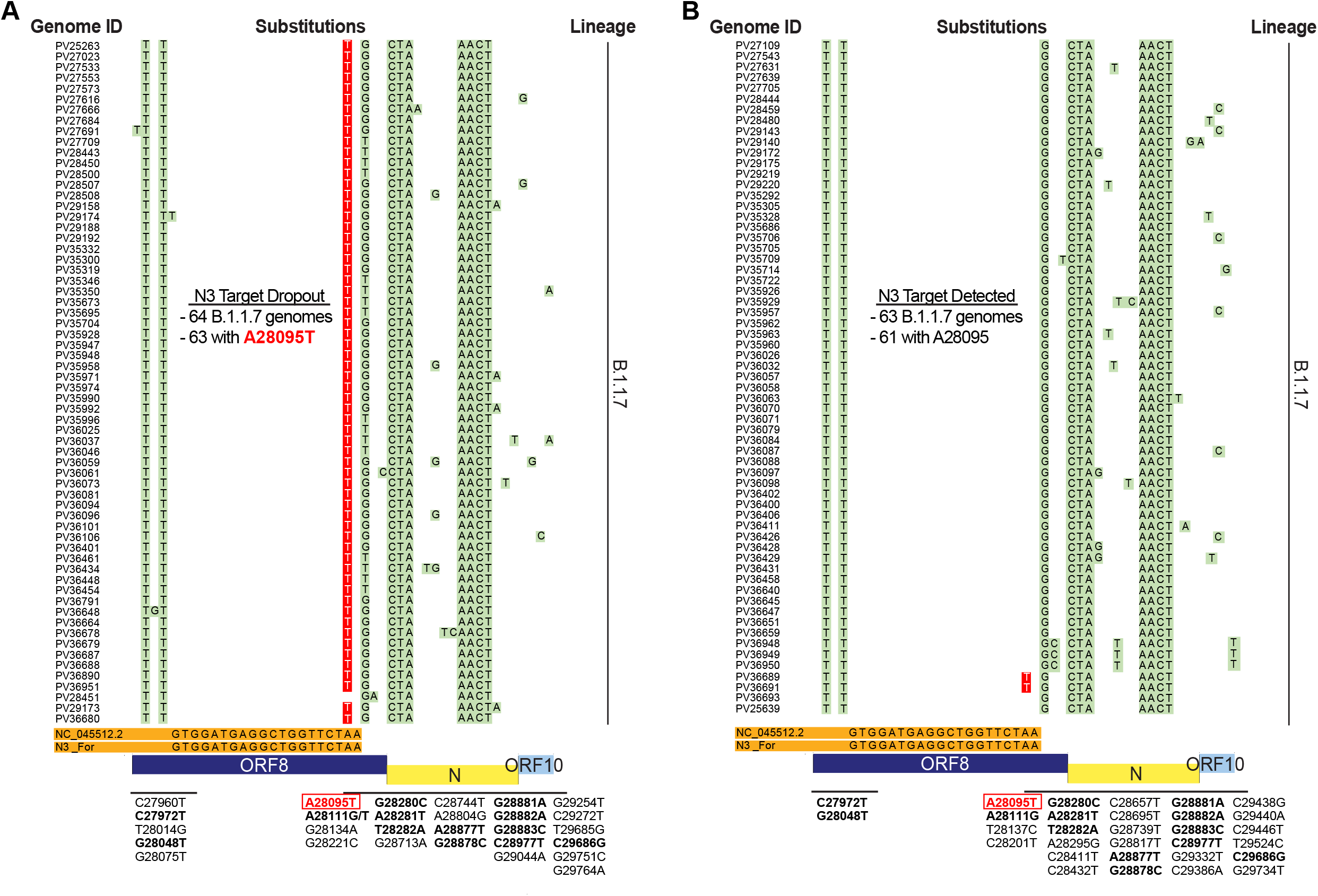
Lineage-specific substitution interferes with SARS-CoV-2 diagnostic target detection. **A:** Alignment of B.1.1.7 genomes associated with N3 target dropout. View is magnified to display mismatches across the N3 forward (For) primer binding site. Sixty-four specimen genomes are indicated by laboratory identifiers (Genome ID) and mismatches to the Wuhan-Hu-1 reference sequence (NC_045512.2) and the N3 For primer (orange) are highlighted in green. Substitutions that correspond with each of the mismatches are annotated below each panel. The lineage specific A28095T substitution that is associated with N3 target dropout is highlighted in red with white typeface font. **B:** Alignment of B.1.1.7 genomes associated with N3 target detection. Sixty-three individual genomes are indicated by laboratory identifiers and mismatches are highlighted and annotated as in **A**. Note for substitutions that are shared across both target results (e.g., dropout and detected), annotations are in boldface font.

Among the other 45 specimens with N3 target dropout, 10 harbored mismatches in the N3 PBS (**Supplemental Figure S2**). Of the 10 genomes, only one (specimen PV36946) had the A28095T substitution, but the sequence recovered was incomplete (42% completeness) and a lineage could not be assigned. The other 9 genomes represented B.1.2 (n = 3), B.1.36.18 (n = 1), B.1.427 (n = 2), B.1.575 (n = 2), and B.1.621 (n = 1) lineages. Among these non-B.1.1.7 genomes, three mismatches were identified at the 1^st^ (G28077T; n = 1), 11^th^ (C28087T; n = 3) and 17^th^ (C28093T; n = 5) bp of the N3 forward PBS which were encoded by viruses from multiple lineages. Of note, these non-B.1.1.7 genomes did not have any other mismatches in the N3 reverse or probe PBS. Furthermore, the two mismatches closest to the 3’ end of the N3 forward primer – C28087T and C28093T – were significantly associated with N3 target dropout (Fisher’s exact, p = 0.0407 and p < 0.0001, respectively), but the mismatch at the first position was not significantly associated with N3 target dropout (Fisher’s exact, p = 0.0914).

## Discussion

Molecular assays for the diagnosis of COVID-19 developed early in the pandemic utilize primers and probes based on conserved regions in the then-available SARS-CoV-2 genome sequences. Now, more than 18 months later, circulating SARS-CoV-2 variants have accumulated numerous nucleotide substitutions in response to evolutionary pressures. These genomic variations can be associated with increased infectivity, transmissibility, and disease pathogenesis ^32–36^, warranting accurate and quick surveillance efforts. However, genome variation can also be a challenge for detection of these variants of interest (VOIs) or VOCs if mismatches to PBSs in diagnostic targets are present. A number of studies have described substitutions in the *ORF1ab, S, E*, and *N* genes that may interfere with specific RT-PCR targets ^1,5,9,11,12,14–16,18^, but these studies have inherent limitations. Several are *in silico* analyses that do not reflect diagnostic performance in the clinical setting ^1,5,16,18^, whereas others do not definitively demonstrate target dropout due to substitutions as they utilize platforms for which primer/probe sequence information is not publicly available ^9,11,14,15^. In addition, the later studies are based on assays that interrogate up to 3 diagnostic targets, and are limited by the number and diversity of viral sequences surveyed over finite timeframes, some prior to the emergence and expansion of VOCs.

In the current study, we describe a robust evaluation of the impact of PBS mismatches on Agena MassARRAY^®^ SARS-CoV-2 Panel target results for over 1,200 specimens over a five-month time period that corresponds with the rapid emergence of viral VOIs/VOCs (December 2020 through April 2021). This large dataset enabled direct correlation of detection of each of five different Agena diagnostic viral targets with genomic sequence data

Additionally, by using publicly available primer/probe sequences to map lineage-specific substitutions, we were able to further evaluate the impact of mismatches on target results and to demonstrate an association between variation in SARS-CoV-2 PBSs and target dropout. Our analysis revealed that several mutations result in N1 and N3 target dropout. Interestingly, although specimens from other lineages harbor mismatches in the N3 target region, we identified a distinct association between the B.1.1.7-associated A28095T substitution and dropout of the N3 diagnostic target on the Agena MassARRAY^®^ SARS-CoV-2 Panel. This finding represents the first description of a lineage-specific substitution that introduces a mismatch to a publicly available primer sequence and yields diagnostic target dropout. This underscores the utility of publicly available sequences to further monitor their diagnostic ability as SARS-CoV-2 continues to evolve and new lineages emerge.

The B.1.1.7 lineage (alpha) has been designated as a variant of concern by the World Health Organization and the US Centers for Diseases Control and Prevention due to its increased transmissibility ^20,33–35,37^. This lineage was first reported in > 1,100 cases in the United Kingdom (UK) on December 14, 2020 ^38^, but it has been estimated to have emerged in September 2020 ^20,35^. Since then, the B.1.1.7 lineage spread rapidly, comprising > 90% of new SARS-CoV-2 infections in the UK by March 2021 ^35^. This lineage also spread globally, including in the US ^39,40^, where recent epidemiological reports indicate B.1.1.7 variants caused > 60% of the new infections as of May 6, 2021 ^41^.

These characteristics further underscore the urgency to update and continually develop robust screening modalities to capture VOCs like B.1.1.7. Sequencing of these variants remains the ‘gold standard’ of surveillance, but not all diagnostic laboratories have the infrastructure or capacity to readily utilize this technology. *S* gene target failure (SGTF) on commercial RT-PCR platforms has been proposed as a screening alternative to detect the S protein deletion H69-V70 (ΔH69/ΔV70) ^14,15,42,43^ which is found in B.1.1.7 and, to a lesser extent, in other circulating lineages (e.g., B.1.525, B.1.620 (NextStrain, build June 23, 2021)) ^15^.

In this study, we describe diagnostic target dropout that can be utilized to promptly identify specimens of interest for whole genome sequencing and variant classification. Notably detection of the B.1.1.7 variant containing the A28095T substitution is associated with the Agena N3 target dropout. This substitution is characteristic of 50% of the circulating B.1.1.7 specimens in our dataset and reflects a unique snapshot of genomic variation occurring within the circulating B.1.1.7 variants in New York City. The A28095T substitution introduces a stop codon in the *ORF8* gene producing a truncated version with potential functional changes in encoded protein (K68stop). Among a dataset of >2 million publicly available viral genomes from global surveillance efforts (GISAID, June 23, 2021), 309,050 genomes harbor this substitution. Nearly all (99.9%) of these A28095T-genomes belong to the B.1.1.7 lineage which, in turn, represent a sub-population (33.9%) of all B.1.1.7 genomes. Therefore, continued diagnostic surveillance of the Agena N3 target dropout and subsequent genomic surveillance can be exploited to monitor the spread of the B.1.1.7 variant as well as other VOCs. Indeed, based on publicly available genomes, other VOCs harbor substitutions that result in mismatches to Agena target PBSs. For example, 73% of delta (B.1.617.2) variant genomes have the non-synonymous substitution, G28916T in the *N* gene (amino acid change, G215C) which introduces a mismatch to the terminal bp of the N2 probe (GISAID, June 23, 2021). Given its position in the probe, this mismatch likely impacts N2 target performance. This warrants further study, particularly as the B.1.617.2 VOC continues to expand globally ^44–46^ since its parent lineage was first identified in India in October 2020 ^47–49^.

An important potential limitation of our study is that target performance can be affected when the quantity of viral nucleic acids in diagnostic specimens is at or near the assay limit of detection, and that the limit of detection varies for different targets. We have demonstrated previously that Agena MassARRAY^®^ target detection is proportional to quantity of viral nucleic acids ^23^. Thus, detection of other targets can be used as a control to evaluate the performance of an individual target, such as N3 target dropout in the setting of the B.1.1.7 A28095T variant. In addition, we have also identified other mutations in this study that are associated with N1 and N3 target dropout. However, unlike the B.1.1.7 A28095T/A genomes in our dataset, these are fewer in number and further comprehensive evaluation is needed to determine the definitive impact on target performance.

Assay platforms that incorporate testing of multiple targets within the virus genome are more likely to retain diagnostic sensitivity as SARS-CoV-2 continues to diversify and new variants emerge. Diagnostic target performance patterns on these redundant platforms have the potential to accommodate unfolding genomic variation in a timely manner, and highlight the potential of diagnostic results to serve as a robust system for detection of these emergent SARS-CoV-2 variants. These qualities demonstrate the importance of these platforms to capture the evolutionary consequences of the ongoing pandemic to inform public health and infection prevention measures.

## Data Availability

SARS-CoV-2 sequencing read data for all study genomes were deposited in GISAID [www.gisaid.org] (accessions pending).

## Code availability

To generate genome sequences, sequencing data were analyzed using a custom reference-based (MN908947.3) pipeline, https://github.com/mjsull/COVID_pipe ^50^. To analyze mismatches to diagnostic target PBSs, genome sequences were analyzed using a custom Unix-code https://github.com/AceM1188/SACOV_primer-probe_analyses ^31^.

## Acknowledgments

We thank the members of MSHS CML, Simon, and van Bakel laboratories for providing any assistance when needed throughout this study. We are grateful for the continuous expert guidance provided by the ISMMS Program for the Protection of Human Subjects (PPHS).

The Research reported in this paper was supported by the National Institutes of Health (NIH) contract number HHSN272201400008C, the NIH Office of Research Infrastructure under award numbers S10OD018522 and S10OD026880, institutional and philanthropic funds (Open Philanthropy Project, #2020-215611), as well as a Robin Chemers Neustein Postdoctoral Fellowship Award (to Dr. Gonzalez-Reiche).

## Author contributions

M.M.H., R.B., P.S., H.A., S.F., A.A., A.E.PM., M.R.G., M.D.N., and E.M.S. provided clinical samples for the study. M.M.H., R.B., P.S., L.C., F.C., H.S., and A.E.PM. accessioned clinical samples. A.S.GR., A.v.d.G., Z.K., B.A., and H.v.B. performed NGS experiments. R.S. provided NGS services. A.S.GR., and A.O. performed genome assembly, data curation and genotyping. M.M.H, L.H.P, and J.D.R. performed alignments and mismatch analyses. M.M.H., R.B., L.H.P, J.D.R., M.R.G., M.D.N., C.C.C., T.E.S., V.S., H.v.B., E.M.S., and A.E.PM analyzed, interpreted, or discussed data. M.M.H., E.M.S., and A.E.PM. wrote the manuscript. M.M.H., E.M.S., and A.E.PM. conceived the study. E.M.S. and A.E.PM. supervised the study. H.v.B., V.S., and E.M.S. raised financial support.

M.M.H. and A.E.PM are the guarantors of this work and, as such, had full access to all of the data in the study and take responsibility for the integrity of the data and the accuracy of the data analysis.

## Competing Interests

Robert Sebra is VP of Technology Development and a stockholder at Sema4, a Mount Sinai Venture. This work, however, was conducted solely at Icahn School of Medicine at Mount Sinai. Otherwise, the authors declare no competing interests.

