## Supplemental Info for "Robust clinical detection of SARS-CoV-2 variants by RT-PCR/MALDI-TOF multi-target approach"

**SUPPLEMENTAL DATA**

**Supplemental Table S1. Agena MassARRAY^®^ target primer and probe sequences**

| **Target** | **Primer/Probe** | **Sequence (5’->3’)** | **SARS-CoV-2 Coordinates (NC_045512.2)** |
| --- | --- | --- | --- |
| N1 | For | AGACGGCATCATATGGGTTG | 28654 - 28673 |
| N1 | Rev | TGAGGAAGTTGTAGCACGATTG | 28761 - 28740 |
| N1 | Probe | GTGCCAATGTGATCTTT | 28716 - 28700 |
| N2 | For | GGGGAACTTCTCCTGCTAGAAT | 28881 - 28902 |
| N2 | Rev | CAGACATTTTGCTCTCAAGCTG | 28979 - 28958 |
| N2 | Probe | GCAAAGCAAGAGCAGCATCACC | 28937 - 28916 |
| N3 | For | GTGGATGAGGCTGGTTCTAA | 28096 - 28077 |
| N3 | Rev | ACTACAAGACTACCCAATTT | 28192 - 28173 |
| N3 | Probe | GAAACTGTATAATTACCGATA | 28138 - 28118 |
| ORF1 | For | AACTGTTGGTCAACAAGACG | 3223 - 3242 |
| ORF1 | Rev | CAATAGTCTGAACAACTGGTGT | 3335 - 3314 |
| ORF1 | Probe | GGTTCAACCTCAATTAG | 3286 - 3302 |
| ORF1AB | For | CCCTGTGGGTTTTACACTTAA | 13342 - 13362 |
| ORF1AB | Rev | ACGATTGTGCATCAGCTGA | 13460 - 13442 |
| ORF1AB | Probe | ATCAACTCCGCGAACCC | 13432 - 13416 |

**Supplemental Table S2. Targets detected for individual specimens.**

| **Specimen ID** | **N1*** | **N2*** | **N3*** | **ORF1A*** | **ORF1AB*** |
| --- | --- | --- | --- | --- | --- |
| PV23213 | D | D | D | D | D |
| PV23214 | D | D | D | D | D |
| PV24080 | D | D | D | D | D |
| PV24082 | D | D | D | D | D |
| PV24084 | D | D | D | D | D |
| PV24085 | D | D | D | D | D |
| PV23212 | D | D | D | D | D |
| PV24087 | D | D | D | D | D |
| PV24088 | D | D | D | D | D |
| PV24092 | D | D | D | D | D |
| PV24049 | D | D | D | D | D |
| PV24050 | D | D | D | D | ND |
| PV24056 | D | D | D | D | D |
| PV24051 | D | D | D | D | D |
| PV24052 | D | D | D | D | D |
| PV24053 | D | D | D | D | D |
| PV24054 | D | D | D | D | D |
| PV24058 | D | D | D | D | D |
| PV24057 | D | D | D | D | D |
| PV24059 | D | D | D | D | D |
| PV24060 | D | D | D | D | D |
| PV24061 | D | D | D | D | D |
| PV24062 | ND | D | D | D | D |
| PV24063 | D | D | D | D | D |
| PV24064 | D | D | D | D | D |
| PV24065 | D | D | D | D | D |
| PV24066 | D | D | D | D | D |
| PV24068 | D | D | D | D | D |
| PV24070 | D | D | D | D | D |
| PV24071 | D | D | D | D | D |
| PV24032 | D | D | D | D | D |
| PV24034 | D | D | D | ND | D |
| PV24036 | D | ND | D | D | D |
| PV24037 | D | D | D | D | D |
| PV24038 | D | D | D | D | D |
| PV24039 | D | D | D | D | D |
| PV24040 | D | D | D | D | D |
| PV24823 | D | D | D | D | D |
| PV24041 | D | D | D | D | D |
| PV24042 | D | D | D | D | D |
| PV24043 | D | D | D | D | D |
| PV24044 | D | D | D | D | D |
| PV24045 | D | D | D | D | D |
| PV24047 | D | D | D | D | D |
| PV24048 | D | D | D | D | D |
| PV24804 | D | D | D | D | D |
| PV24814 | D | D | D | D | D |
| PV24813 | D | D | D | D | D |
| PV24810 | D | D | D | D | D |
| PV24818 | D | D | D | D | D |
| PV24820 | D | D | D | D | D |
| PV24817 | D | D | D | D | D |
| PV24815 | D | D | D | D | D |
| PV24812 | D | D | D | D | D |
| PV24807 | D | D | D | D | D |
| PV24816 | D | D | ND | D | ND |
| PV24803 | D | D | D | D | D |
| PV24808 | D | D | D | D | D |
| PV24819 | D | D | D | D | D |
| PV24809 | D | D | D | D | D |
| PV24806 | D | D | D | D | D |
| PV24821 | D | D | D | D | D |
| PV24832 | D | D | D | D | D |
| PV24847 | D | D | D | D | D |
| PV24830 | D | D | D | D | D |
| PV24825 | D | D | ND | D | D |
| PV24837 | D | D | D | D | D |
| PV25218 | D | D | D | D | D |
| PV25199 | D | D | D | D | D |
| PV25189 | D | D | D | D | D |
| PV25212 | D | D | D | D | D |
| PV24844 | D | D | D | D | D |
| PV25205 | D | D | ND | D | D |
| PV24836 | D | D | D | D | D |
| PV24829 | D | D | D | D | D |
| PV24845 | D | D | D | D | D |
| PV24834 | D | D | D | D | D |
| PV24842 | D | D | D | D | D |
| PV24841 | D | D | D | D | D |
| PV24840 | D | D | D | D | D |
| PV24843 | D | D | D | D | D |
| PV24873 | D | ND | ND | D | ND |
| PV24874 | D | D | D | D | D |
| PV24876 | D | D | ND | D | D |
| PV24879 | D | D | D | D | D |
| PV24883 | D | D | ND | D | D |
| PV24893 | D | D | D | D | D |
| PV24887 | D | D | D | D | D |
| PV24888 | D | D | D | D | D |
| PV24889 | D | D | D | D | D |
| PV24885 | D | D | D | D | D |
| PV24890 | D | D | D | D | D |
| PV24891 | D | D | ND | D | D |
| PV24892 | D | D | D | D | D |
| PV24886 | D | D | D | D | D |
| PV24899 | D | D | D | D | D |
| PV24926 | D | ND | ND | D | ND |
| PV24901 | D | D | D | D | D |
| PV24902 | D | D | D | D | D |
| PV24913 | D | D | D | D | D |
| PV24927 | D | D | D | D | D |
| PV24895 | D | D | D | D | D |
| PV24908 | D | D | D | D | D |
| PV24935 | D | D | D | D | D |
| PV24910 | D | ND | ND | D | ND |
| PV24915 | D | D | ND | D | D |
| PV24919 | D | D | D | D | D |
| PV24933 | D | D | D | D | D |
| PV24928 | D | D | D | D | D |
| PV24903 | D | D | D | D | D |
| PV23447 | D | D | D | D | D |
| PV24912 | D | D | D | D | ND |
| PV24934 | D | D | D | D | D |
| PV24914 | D | D | D | D | D |
| PV24916 | D | D | D | D | D |
| PV24905 | D | D | D | D | D |
| PV24918 | D | D | D | D | D |
| PV24920 | D | D | D | D | D |
| PV25192 | D | D | D | D | D |
| PV25206 | D | D | D | D | D |
| PV25220 | D | D | D | D | D |
| PV25198 | D | D | D | D | D |
| PV25194 | D | D | D | D | D |
| PV25188 | D | D | D | D | D |
| PV25217 | D | D | D | D | D |
| PV25216 | D | D | D | D | D |
| PV25187 | D | D | D | D | D |
| PV25219 | D | D | D | D | D |
| PV25238 | D | D | D | D | D |
| PV25234 | D | D | D | D | D |
| PV25213 | D | D | D | D | D |
| PV25229 | D | D | D | D | D |
| PV25227 | D | D | D | D | D |
| PV25235 | D | D | D | D | D |
| PV25240 | D | D | D | D | D |
| PV25242 | D | ND | D | D | D |
| PV25241 | D | D | D | D | D |
| PV25245 | D | D | D | D | D |
| PV25246 | D | D | D | D | D |
| PV25224 | D | D | D | D | D |
| PV25221 | D | D | D | D | D |
| PV25247 | D | D | D | D | D |
| PV25256 | D | D | D | D | D |
| PV25231 | D | D | D | D | D |
| PV25237 | D | D | D | D | D |
| PV25257 | D | D | ND | D | D |
| PV25228 | D | D | D | D | D |
| PV25254 | D | D | D | D | D |
| PV25244 | D | D | D | D | D |
| PV25252 | D | D | D | D | D |
| PV25250 | D | D | D | D | D |
| PV25236 | D | D | D | D | D |
| PV25248 | D | D | D | D | D |
| PV25253 | D | D | ND | D | D |
| PV25233 | D | D | D | D | D |
| PV25226 | D | D | D | D | ND |
| PV25223 | D | D | D | D | D |
| PV25230 | D | D | D | D | ND |
| PV25249 | D | D | D | D | D |
| PV25618 | D | D | D | D | D |
| PV25619 | D | ND | ND | D | ND |
| PV25620 | D | D | D | D | D |
| PV25647 | D | D | D | D | D |
| PV25623 | D | D | D | D | D |
| PV25624 | D | D | D | D | D |
| PV25282 | D | D | D | D | D |
| PV25625 | D | D | D | D | D |
| PV25622 | D | D | D | D | D |
| PV25626 | D | D | D | D | D |
| PV25627 | D | D | D | D | D |
| PV25628 | D | D | D | D | D |
| PV25629 | D | D | ND | D | D |
| PV25630 | D | D | D | D | D |
| PV25643 | D | D | D | D | D |
| PV25644 | D | D | D | D | D |
| PV25631 | D | ND | ND | D | ND |
| PV25635 | D | D | D | D | D |
| PV25636 | D | D | D | D | D |
| PV25637 | D | D | D | D | D |
| PV25645 | D | D | D | D | D |
| PV25633 | D | D | D | D | D |
| PV25639 | D | D | D | ND | ND |
| PV25640 | D | D | D | D | D |
| PV25641 | D | D | D | D | D |
| PV25646 | D | D | D | D | D |
| PV25634 | D | D | D | D | D |
| PV25642 | D | D | D | D | D |
| PV25650 | D | D | D | D | D |
| PV25653 | D | D | D | D | D |
| PV25651 | D | D | D | D | D |
| PV25652 | D | D | D | D | D |
| PV25298 | D | D | D | D | D |
| PV25671 | D | D | D | D | D |
| PV25654 | D | D | D | D | D |
| PV25658 | D | D | D | D | D |
| PV25659 | D | D | D | D | D |
| PV25664 | D | D | D | D | D |
| PV25665 | D | D | D | D | D |
| PV25666 | D | D | D | D | D |
| PV25667 | D | ND | D | D | D |
| PV25672 | D | D | D | D | D |
| PV25673 | D | D | D | D | D |
| PV25668 | D | D | D | D | ND |
| PV25669 | D | D | D | D | ND |
| PV25674 | D | D | D | D | D |
| PV25670 | D | D | D | D | D |
| PV25675 | D | D | D | D | D |
| PV25676 | D | D | D | D | D |
| PV25683 | D | D | D | D | D |
| PV25684 | D | D | D | D | D |
| PV25685 | D | D | D | D | D |
| PV25677 | D | D | D | D | D |
| PV25678 | D | D | D | D | D |
| PV25679 | D | D | D | D | D |
| PV25680 | D | D | D | D | D |
| PV25681 | D | D | D | D | ND |
| PV25682 | D | D | D | D | D |
| PV25787 | D | D | D | D | D |
| PV25788 | D | D | D | D | D |
| PV25790 | D | D | D | D | D |
| PV25789 | D | D | D | D | D |
| PV25791 | D | D | D | D | D |
| PV25792 | D | D | D | D | D |
| PV25795 | D | D | D | D | D |
| PV25794 | D | D | D | D | D |
| PV25024 | D | D | D | D | D |
| PV25801 | D | D | D | D | D |
| PV25802 | D | D | D | D | D |
| PV25803 | D | D | D | D | D |
| PV25796 | D | D | D | D | D |
| PV25804 | D | D | D | D | D |
| PV25797 | D | D | D | D | D |
| PV25798 | D | D | D | D | D |
| PV25805 | D | D | D | D | D |
| PV25799 | D | D | D | D | D |
| PV25806 | D | D | D | D | D |
| PV25808 | D | D | D | D | D |
| PV25809 | D | D | D | D | D |
| PV25810 | D | D | D | D | D |
| PV25811 | D | D | D | D | D |
| PV25812 | D | D | D | D | D |
| PV25814 | D | D | D | D | D |
| PV25821 | D | D | D | D | D |
| PV25815 | D | D | D | D | D |
| PV25816 | D | D | D | D | D |
| PV25817 | D | D | D | D | D |
| PV25822 | D | D | D | D | D |
| PV25818 | D | D | D | D | D |
| PV25823 | D | D | D | D | D |
| PV25819 | D | D | D | D | D |
| PV25824 | D | D | D | D | D |
| PV25820 | D | D | D | D | ND |
| PV25827 | D | D | D | D | D |
| PV25825 | D | D | D | D | D |
| PV25828 | D | D | D | D | D |
| PV25829 | D | D | D | D | D |
| PV25830 | D | D | D | D | D |
| PV25831 | D | D | D | D | D |
| PV25826 | D | D | D | D | D |
| PV25835 | D | D | D | D | D |
| PV25836 | D | D | D | D | D |
| PV25837 | D | D | D | D | D |
| PV25839 | D | D | D | D | D |
| PV25840 | D | D | D | D | D |
| PV25841 | D | D | D | D | D |
| PV25842 | D | D | D | D | D |
| PV25832 | D | D | D | D | D |
| PV25833 | D | D | D | D | D |
| PV25834 | D | D | D | D | D |
| PV25843 | D | D | D | D | D |
| PV25846 | D | ND | D | D | D |
| PV25847 | D | D | D | D | D |
| PV25849 | D | D | D | D | ND |
| PV25851 | D | D | D | D | D |
| PV25844 | D | D | D | D | ND |
| PV25850 | D | D | D | D | D |
| PV25853 | D | D | D | D | D |
| PV25858 | D | D | D | D | D |
| PV25879 | D | D | D | D | D |
| PV25880 | D | D | D | D | D |
| PV25875 | D | D | D | D | D |
| PV25876 | D | D | D | D | D |
| PV25859 | D | D | D | D | D |
| PV25860 | D | D | D | D | D |
| PV25861 | D | D | D | D | ND |
| PV25862 | D | D | D | D | D |
| PV25852 | D | D | D | D | D |
| PV25857 | D | D | D | D | D |
| PV25864 | D | D | ND | D | ND |
| PV25856 | D | D | D | D | D |
| PV25865 | D | D | D | D | D |
| PV25866 | D | D | D | D | D |
| PV25869 | D | D | D | D | D |
| PV25877 | D | D | D | D | ND |
| PV25873 | D | D | D | D | D |
| PV25881 | D | D | ND | ND | ND |
| PV25971 | D | D | D | D | D |
| PV25867 | D | D | D | D | D |
| PV25870 | D | D | D | D | D |
| PV25871 | D | D | D | D | D |
| PV25972 | D | D | D | D | D |
| PV25868 | D | D | D | D | D |
| PV25878 | D | D | D | D | D |
| PV25973 | D | D | D | D | D |
| PV25974 | D | D | D | D | D |
| PV25975 | D | D | D | D | D |
| PV25978 | D | D | D | D | D |
| PV25982 | D | D | D | D | D |
| PV25983 | D | D | D | D | D |
| PV25984 | D | D | D | D | ND |
| PV25976 | D | D | D | D | D |
| PV25979 | D | D | D | D | ND |
| PV25985 | D | D | D | D | D |
| PV25986 | D | D | D | D | D |
| PV25980 | D | D | D | D | D |
| PV25981 | D | D | D | D | D |
| PV25990 | D | D | D | D | D |
| PV25988 | D | D | D | D | D |
| PV25987 | D | D | D | D | D |
| PV25989 | D | D | D | D | ND |
| PV25992 | D | D | D | D | D |
| PV25991 | D | D | D | D | ND |
| PV25993 | D | D | D | D | D |
| PV25994 | D | D | ND | D | D |
| PV25995 | D | D | D | D | D |
| PV25996 | D | D | D | D | D |
| PV25997 | D | D | D | D | D |
| PV26004 | D | D | ND | D | D |
| PV26005 | D | D | D | D | D |
| PV26007 | D | D | D | D | D |
| PV26008 | D | D | D | D | D |
| PV26002 | D | D | D | D | D |
| PV26009 | D | D | D | D | D |
| PV26011 | D | D | D | D | D |
| PV26012 | D | D | D | D | D |
| PV26013 | D | D | D | D | D |
| PV26014 | D | D | D | D | D |
| PV26000 | D | ND | D | D | ND |
| PV26010 | D | D | D | D | D |
| PV26001 | D | D | D | D | D |
| PV26003 | D | D | D | D | D |
| PV26015 | D | D | D | D | D |
| PV26016 | ND | D | D | D | D |
| PV26017 | ND | D | D | D | D |
| PV26018 | D | D | D | D | D |
| PV25281 | D | D | ND | D | D |
| PV25296 | D | D | ND | D | D |
| PV25301 | D | D | D | D | D |
| PV25264 | D | D | D | D | D |
| PV25260 | D | D | D | D | D |
| PV26020 | D | D | D | D | D |
| PV26021 | D | D | D | D | D |
| PV26022 | D | D | D | D | D |
| PV26023 | D | D | D | D | D |
| PV25263 | D | D | ND | D | D |
| PV25284 | D | D | D | D | D |
| PV25265 | D | D | D | D | D |
| PV25266 | D | D | D | D | ND |
| PV25259 | D | D | D | D | D |
| PV25297 | D | ND | D | D | ND |
| PV25280 | D | D | D | D | D |
| PV25271 | D | D | D | D | D |
| PV25279 | D | D | D | D | D |
| PV25294 | D | D | D | D | D |
| PV25300 | D | D | D | D | D |
| PV25288 | D | D | D | D | D |
| PV25293 | D | D | D | D | D |
| PV25269 | D | D | D | D | D |
| PV25278 | D | D | D | D | D |
| PV25289 | D | D | D | D | D |
| PV25275 | D | D | D | D | D |
| PV25274 | D | D | D | D | D |
| PV25286 | D | D | D | D | D |
| PV25273 | D | D | D | D | D |
| PV25302 | D | D | D | D | D |
| PV25295 | D | D | D | D | D |
| PV25287 | D | D | D | D | D |
| PV25270 | D | D | D | D | D |
| PV25285 | D | D | D | D | D |
| PV25283 | D | D | ND | D | D |
| PV25268 | D | D | D | D | D |
| PV25272 | D | D | ND | D | D |
| PV25332 | D | D | D | D | D |
| PV25346 | D | ND | D | D | ND |
| PV25339 | D | D | D | D | D |
| PV25347 | D | D | D | D | D |
| PV25340 | D | D | D | D | D |
| PV25333 | D | D | D | D | D |
| PV25334 | D | D | D | D | D |
| PV25335 | D | D | D | D | D |
| PV25348 | D | D | D | D | D |
| PV25342 | D | D | D | D | D |
| PV25336 | D | D | D | D | D |
| PV25337 | ND | D | D | D | D |
| PV25345 | D | D | D | D | D |
| PV25351 | D | D | D | D | D |
| PV25355 | D | D | D | D | D |
| PV25356 | D | D | D | D | D |
| PV25357 | D | D | D | D | ND |
| PV25358 | D | D | D | D | D |
| PV25352 | D | D | D | D | ND |
| PV25353 | D | D | D | D | D |
| PV25354 | D | D | D | D | D |
| PV25361 | D | D | D | D | D |
| PV26297 | D | D | D | D | D |
| PV26282 | D | D | D | D | D |
| PV26284 | D | D | D | D | D |
| PV26285 | D | D | D | D | D |
| PV26286 | D | D | D | D | D |
| PV26287 | D | D | D | D | D |
| PV26298 | D | D | D | D | D |
| PV26288 | D | D | D | D | D |
| PV26301 | D | D | D | D | ND |
| PV26329 | D | D | ND | D | D |
| PV26315 | D | D | D | D | D |
| PV26316 | D | D | D | D | D |
| PV26319 | D | ND | D | D | ND |
| PV26320 | D | D | D | D | D |
| PV26332 | D | D | D | D | D |
| PV26335 | D | D | D | D | D |
| PV26330 | D | D | D | D | D |
| PV26331 | D | D | D | D | D |
| PV26345 | D | D | D | D | D |
| PV26347 | D | D | D | D | D |
| PV26349 | D | D | D | D | D |
| PV26351 | D | D | D | D | D |
| PV26352 | D | D | D | D | D |
| PV26358 | D | D | D | D | D |
| PV26359 | D | D | D | D | D |
| PV26360 | D | D | D | D | D |
| PV26361 | D | D | D | D | D |
| PV26362 | D | D | D | D | D |
| PV26380 | D | D | ND | D | D |
| PV26374 | D | D | D | D | D |
| PV26363 | D | D | D | D | D |
| PV26364 | D | D | D | D | D |
| PV26376 | D | D | D | D | D |
| PV26377 | D | D | ND | D | D |
| PV26381 | D | D | D | D | D |
| PV25649 | D | D | D | D | ND |
| PV26392 | D | D | D | D | D |
| PV26393 | D | D | D | D | D |
| PV26394 | D | D | D | D | D |
| PV26395 | D | D | ND | D | D |
| PV26397 | D | D | D | D | D |
| PV26406 | ND | D | D | D | D |
| PV26407 | D | D | D | D | D |
| PV26409 | D | D | D | D | D |
| PV26410 | D | D | D | D | D |
| PV26411 | D | D | D | D | D |
| PV26412 | D | D | D | D | D |
| PV26413 | D | D | D | D | D |
| PV26422 | D | D | D | D | D |
| PV26425 | D | D | D | D | D |
| PV26426 | D | D | D | D | D |
| PV26429 | D | ND | ND | D | ND |
| PV26502 | D | D | D | D | D |
| PV26488 | D | D | D | D | D |
| PV26482 | D | ND | D | D | ND |
| PV26483 | D | ND | D | D | D |
| PV26504 | D | ND | D | D | ND |
| PV26444 | D | D | D | D | D |
| PV26535 | D | D | D | D | D |
| PV26485 | D | ND | ND | D | ND |
| PV26536 | D | D | D | D | D |
| PV26518 | D | D | ND | D | D |
| PV26515 | D | D | D | D | D |
| PV26519 | D | D | D | D | D |
| PV26498 | D | D | D | D | D |
| PV26545 | D | D | D | D | ND |
| PV26071 | D | D | D | D | D |
| PV26520 | D | D | D | D | D |
| PV26578 | D | D | D | D | D |
| PV26579 | D | D | D | D | D |
| PV26516 | D | D | D | D | D |
| PV26499 | D | D | D | D | D |
| PV26513 | D | D | D | D | D |
| PV26514 | D | D | D | D | D |
| PV26517 | D | D | D | D | D |
| PV26529 | D | D | D | D | D |
| PV26501 | D | D | D | D | D |
| PV26068 | D | D | D | D | D |
| PV26546 | D | D | D | D | D |
| PV26530 | D | ND | D | D | D |
| PV26531 | ND | D | D | D | D |
| PV26552 | D | D | D | D | D |
| PV26561 | D | D | D | D | D |
| PV26562 | D | D | D | D | D |
| PV26563 | D | D | ND | D | D |
| PV26550 | D | D | D | D | D |
| PV26551 | D | D | D | D | D |
| PV26440 | D | D | D | D | D |
| PV26441 | D | D | D | D | D |
| PV26442 | D | D | D | D | D |
| PV26564 | D | D | D | D | D |
| PV26565 | D | D | D | D | D |
| PV26566 | D | D | D | D | ND |
| PV26445 | D | D | D | D | D |
| PV26567 | D | D | D | D | D |
| PV26574 | D | D | D | D | D |
| PV26576 | D | D | D | D | D |
| PV26577 | D | D | D | D | D |
| PV26581 | D | D | D | D | ND |
| PV26443 | D | D | D | D | D |
| PV26590 | D | D | D | D | D |
| PV26591 | D | D | D | D | D |
| PV26592 | D | D | D | D | ND |
| PV26593 | D | D | D | D | ND |
| PV26594 | D | D | ND | D | ND |
| PV26595 | D | D | D | D | D |
| PV26596 | D | D | D | D | D |
| PV26597 | D | D | D | D | D |
| PV26606 | D | D | D | D | ND |
| PV26608 | D | D | D | D | D |
| PV26613 | D | D | D | D | D |
| PV26609 | D | D | D | D | D |
| PV26610 | D | D | D | D | ND |
| PV26607 | D | D | D | D | D |
| PV26622 | D | D | D | D | D |
| PV26624 | D | D | D | D | D |
| PV26625 | D | D | D | D | D |
| PV26627 | D | D | D | D | D |
| PV26629 | D | D | D | D | D |
| PV26611 | D | D | ND | D | D |
| PV26642 | D | D | D | D | D |
| PV26643 | D | D | D | D | D |
| PV26644 | D | D | D | D | D |
| PV26639 | D | D | D | D | D |
| PV26640 | D | D | D | D | D |
| PV26655 | D | D | D | D | D |
| PV26656 | D | ND | D | D | ND |
| PV26660 | D | D | D | D | D |
| PV26933 | D | D | D | D | D |
| PV26934 | D | ND | D | D | ND |
| PV26935 | D | ND | ND | D | ND |
| PV26936 | D | D | D | D | D |
| PV26939 | D | D | D | D | D |
| PV26940 | D | D | D | D | D |
| PV26947 | D | D | D | D | D |
| PV26937 | D | D | D | D | ND |
| PV26950 | D | D | D | D | ND |
| PV26944 | D | D | D | D | D |
| PV26945 | D | D | D | D | D |
| PV26941 | D | D | D | D | ND |
| PV26942 | D | D | D | D | ND |
| PV26938 | D | D | D | D | D |
| PV26943 | D | D | D | D | D |
| PV26956 | D | D | D | D | D |
| PV26946 | D | D | D | D | D |
| PV26951 | ND | D | ND | D | ND |
| PV26948 | D | D | ND | ND | ND |
| PV26967 | D | D | D | D | ND |
| PV26952 | D | D | D | D | ND |
| PV26955 | D | ND | ND | D | ND |
| PV26953 | D | D | ND | ND | ND |
| PV26949 | D | D | D | D | D |
| PV26968 | D | ND | D | D | ND |
| PV26957 | D | D | D | D | ND |
| PV26958 | D | D | D | D | ND |
| PV26959 | D | D | D | D | D |
| PV26960 | D | D | D | D | ND |
| PV26961 | D | D | ND | ND | ND |
| PV26969 | D | D | ND | D | ND |
| PV26962 | D | D | D | D | D |
| PV26963 | D | D | D | D | D |
| PV26964 | D | D | D | D | ND |
| PV26965 | D | D | D | D | D |
| PV26966 | D | D | D | D | D |
| PV26989 | D | ND | ND | D | ND |
| PV26978 | D | D | D | D | D |
| PV26979 | D | D | ND | ND | ND |
| PV26970 | D | D | D | D | D |
| PV26980 | D | ND | ND | D | ND |
| PV26971 | D | D | D | D | ND |
| PV26972 | D | D | D | D | ND |
| PV26981 | D | D | D | D | D |
| PV26982 | D | D | ND | ND | ND |
| PV26973 | D | D | D | D | D |
| PV26975 | D | D | D | D | D |
| PV26976 | D | D | D | D | D |
| PV26977 | D | D | D | D | D |
| PV26983 | D | D | D | D | D |
| PV26984 | D | D | D | D | ND |
| PV26985 | D | D | D | D | D |
| PV26974 | D | D | D | D | D |
| PV26987 | D | D | D | D | D |
| PV26990 | D | D | D | D | D |
| PV26988 | D | D | D | D | D |
| PV26991 | D | D | ND | D | ND |
| PV26992 | D | ND | D | D | D |
| PV26993 | D | ND | D | D | D |
| PV26994 | D | D | D | D | D |
| PV26995 | D | D | D | D | D |
| PV26996 | D | ND | D | D | ND |
| PV26997 | D | D | D | D | D |
| PV26998 | D | D | D | D | D |
| PV26999 | D | D | D | D | D |
| PV26986 | D | D | D | D | D |
| PV27000 | D | D | D | D | D |
| PV27001 | D | D | ND | D | ND |
| PV27002 | D | D | D | D | D |
| PV27003 | D | D | D | D | D |
| PV27004 | D | D | D | D | D |
| PV27010 | D | D | D | D | ND |
| PV27005 | D | D | D | D | D |
| PV27006 | D | ND | ND | D | ND |
| PV27007 | D | D | D | D | D |
| PV27008 | D | D | ND | D | ND |
| PV27009 | D | D | D | D | D |
| PV27023 | D | D | ND | D | D |
| PV27011 | D | ND | D | D | ND |
| PV27018 | D | D | D | D | D |
| PV27012 | D | D | D | D | D |
| PV27013 | D | D | D | D | ND |
| PV27014 | D | D | D | D | D |
| PV27019 | D | D | D | D | D |
| PV27015 | D | D | D | D | D |
| PV27020 | D | D | D | D | D |
| PV27021 | ND | D | D | D | D |
| PV27024 | D | D | D | D | D |
| PV27016 | D | D | D | D | D |
| PV27029 | D | D | D | D | D |
| PV27025 | D | ND | D | D | ND |
| PV27030 | D | D | D | D | D |
| PV27031 | D | D | D | D | D |
| PV27022 | D | D | ND | ND | ND |
| PV27032 | D | D | D | D | D |
| PV27033 | D | D | D | D | D |
| PV27034 | D | D | D | D | D |
| PV27035 | D | D | D | D | D |
| PV27036 | D | D | D | D | D |
| PV27037 | ND | D | D | D | D |
| PV27038 | D | D | D | D | D |
| PV27040 | D | D | D | D | D |
| PV27043 | D | D | D | ND | D |
| PV27041 | D | D | D | D | D |
| PV27039 | D | D | D | D | D |
| PV27042 | D | D | D | D | D |
| PV27047 | D | D | D | D | ND |
| PV27050 | D | D | D | D | D |
| PV27051 | D | D | D | D | D |
| PV27048 | D | D | D | D | D |
| PV27045 | D | D | D | D | D |
| PV27046 | D | ND | D | D | D |
| PV27053 | D | D | D | D | D |
| PV27054 | D | D | D | D | D |
| PV27055 | D | D | D | D | D |
| PV27056 | D | D | D | D | D |
| PV27064 | D | D | D | D | D |
| PV27067 | D | D | D | D | D |
| PV27065 | D | D | D | D | ND |
| PV27058 | D | D | D | D | ND |
| PV27066 | D | D | D | D | D |
| PV27063 | D | D | D | D | ND |
| PV27072 | D | ND | D | D | ND |
| PV27075 | D | D | D | D | D |
| PV27079 | D | D | D | D | D |
| PV27081 | D | D | D | D | D |
| PV27082 | D | D | D | D | D |
| PV27085 | D | D | D | D | D |
| PV27086 | D | D | D | D | D |
| PV27087 | D | D | D | D | D |
| PV27090 | D | D | D | D | D |
| PV27092 | D | D | D | D | D |
| PV27093 | D | D | D | D | D |
| PV27094 | D | D | D | D | D |
| PV27098 | D | D | D | D | D |
| PV27099 | D | D | D | D | D |
| PV27100 | D | D | ND | D | ND |
| PV27101 | D | D | D | D | D |
| PV27104 | D | D | D | D | D |
| PV27103 | D | D | D | D | D |
| PV27105 | D | D | D | D | D |
| PV27107 | D | D | D | D | D |
| PV27108 | D | D | D | D | D |
| PV27109 | D | D | D | D | ND |
| PV27111 | D | D | D | D | D |
| PV27115 | D | D | D | D | D |
| PV27116 | D | D | D | D | D |
| PV27112 | D | D | D | D | D |
| PV27113 | D | ND | D | D | ND |
| PV27114 | D | ND | D | D | D |
| PV27120 | D | D | D | D | D |
| PV27524 | D | D | D | D | D |
| PV27525 | D | D | D | D | D |
| PV27529 | D | D | D | D | D |
| PV27531 | D | D | D | D | D |
| PV27526 | D | D | D | D | D |
| PV27528 | D | D | D | D | D |
| PV27532 | D | D | D | D | D |
| PV27535 | D | D | D | D | D |
| PV27536 | D | ND | D | ND | ND |
| PV27527 | D | D | D | D | D |
| PV27533 | D | D | ND | D | D |
| PV27542 | D | D | D | D | ND |
| PV27539 | D | D | D | D | D |
| PV27534 | D | D | D | D | D |
| PV27540 | D | D | D | D | D |
| PV27543 | D | D | D | D | D |
| PV27544 | D | D | D | D | D |
| PV27545 | D | D | D | D | D |
| PV27541 | D | D | D | D | D |
| PV27547 | D | ND | ND | D | ND |
| PV27548 | D | D | D | D | D |
| PV27546 | D | D | D | D | D |
| PV27550 | D | D | D | D | D |
| PV27553 | D | D | ND | D | D |
| PV27549 | D | D | D | D | D |
| PV27555 | D | D | D | D | D |
| PV27557 | D | D | D | D | D |
| PV27554 | D | D | D | D | ND |
| PV27558 | D | D | D | D | D |
| PV27559 | D | D | D | D | D |
| PV27560 | D | D | D | D | ND |
| PV27562 | D | D | D | D | D |
| PV27565 | D | D | D | D | D |
| PV27566 | D | D | D | D | D |
| PV27567 | D | D | D | D | D |
| PV27568 | D | D | D | D | D |
| PV27563 | D | D | D | D | D |
| PV27569 | D | D | D | D | D |
| PV27570 | D | D | D | D | D |
| PV27571 | D | D | D | D | D |
| PV27572 | D | D | D | D | D |
| PV27573 | D | D | ND | D | D |
| PV27575 | D | D | D | D | D |
| PV27576 | D | ND | D | D | ND |
| PV27601 | D | D | D | D | D |
| PV27581 | D | D | D | D | D |
| PV27577 | D | D | D | D | ND |
| PV27579 | D | D | D | D | ND |
| PV27580 | D | D | D | D | D |
| PV27582 | D | D | D | D | D |
| PV27595 | D | D | D | D | D |
| PV27583 | D | D | D | D | D |
| PV27586 | D | D | D | D | D |
| PV27587 | D | D | D | D | D |
| PV27588 | D | D | D | D | ND |
| PV27584 | D | D | D | D | D |
| PV27603 | D | D | D | D | ND |
| PV27589 | D | D | D | D | ND |
| PV27590 | D | D | D | D | D |
| PV27593 | D | D | D | D | D |
| PV27597 | D | D | D | D | D |
| PV27598 | D | D | D | D | D |
| PV27605 | D | D | D | D | D |
| PV27612 | D | D | D | D | D |
| PV27613 | D | D | D | D | D |
| PV27614 | D | D | D | D | D |
| PV27607 | D | D | D | D | D |
| PV27608 | D | D | D | D | D |
| PV27609 | D | D | D | D | D |
| PV27610 | D | D | D | D | D |
| PV27621 | D | D | D | D | D |
| PV27622 | D | D | D | D | D |
| PV27623 | D | D | D | D | D |
| PV27624 | D | D | ND | D | D |
| PV27615 | D | D | D | D | D |
| PV27627 | D | D | D | D | ND |
| PV27628 | D | D | D | D | ND |
| PV27630 | D | D | D | D | D |
| PV27632 | D | D | D | D | ND |
| PV27616 | D | D | ND | D | D |
| PV27617 | D | ND | ND | D | ND |
| PV27618 | D | D | D | D | D |
| PV27619 | D | D | ND | D | ND |
| PV27620 | D | D | ND | D | D |
| PV27631 | D | D | D | D | ND |
| PV27633 | D | D | D | D | D |
| PV27635 | D | D | D | D | D |
| PV27643 | D | D | D | D | D |
| PV27636 | D | D | D | D | D |
| PV27638 | D | D | D | D | D |
| PV27639 | D | ND | D | D | ND |
| PV27644 | D | D | D | D | D |
| PV27645 | D | D | ND | ND | ND |
| PV27641 | D | D | ND | D | D |
| PV27648 | D | D | D | D | D |
| PV27646 | D | D | D | D | ND |
| PV27647 | D | ND | D | D | ND |
| PV27650 | D | D | D | D | ND |
| PV27651 | D | D | D | D | D |
| PV27656 | D | D | D | D | D |
| PV27657 | D | D | D | D | D |
| PV27658 | D | D | D | D | D |
| PV27653 | D | D | D | D | D |
| PV27654 | D | D | D | D | D |
| PV27655 | D | D | D | D | D |
| PV27659 | D | D | D | D | D |
| PV27666 | ND | D | ND | D | D |
| PV27663 | D | D | ND | D | ND |
| PV27664 | D | D | ND | ND | ND |
| PV27665 | D | D | D | D | D |
| PV27660 | D | D | D | D | D |
| PV27661 | D | D | D | D | D |
| PV27662 | D | D | D | D | D |
| PV27670 | D | ND | D | D | ND |
| PV27671 | D | D | D | D | D |
| PV27673 | D | D | ND | D | D |
| PV27672 | D | D | D | D | D |
| PV27674 | D | D | D | D | D |
| PV27675 | D | D | D | D | D |
| PV27677 | D | D | D | D | D |
| PV27676 | D | D | D | D | D |
| PV27679 | D | D | D | D | ND |
| PV27682 | D | D | D | D | D |
| PV27686 | D | D | D | D | ND |
| PV27680 | D | D | D | D | ND |
| PV27687 | D | D | D | D | D |
| PV27683 | D | D | D | D | D |
| PV27688 | D | D | D | D | D |
| PV27684 | D | D | ND | D | D |
| PV27685 | D | D | D | D | D |
| PV27692 | D | D | D | D | D |
| PV27690 | D | D | D | D | D |
| PV27691 | D | D | ND | D | D |
| PV27695 | D | ND | D | D | ND |
| PV27696 | D | D | D | D | D |
| PV27698 | D | D | D | D | D |
| PV27700 | ND | D | D | D | D |
| PV27701 | D | D | D | D | D |
| PV27702 | D | D | D | D | D |
| PV27703 | D | D | D | D | ND |
| PV27706 | D | D | D | D | D |
| PV27704 | D | D | D | D | D |
| PV27705 | D | D | D | D | D |
| PV27708 | D | D | D | D | D |
| PV27709 | D | D | ND | D | D |
| PV27707 | D | D | D | D | ND |
| PV27710 | D | D | D | D | D |
| PV27711 | D | D | D | D | D |
| PV28420 | D | D | D | D | D |
| PV27712 | D | D | D | D | D |
| PV28417 | D | D | D | D | D |
| PV28419 | D | D | D | D | D |
| PV28421 | D | ND | D | D | D |
| PV28422 | D | D | D | D | D |
| PV28430 | D | D | D | D | D |
| PV28425 | D | D | D | D | D |
| PV28428 | D | D | D | D | D |
| PV28431 | D | D | D | D | ND |
| PV28432 | D | D | D | D | D |
| PV28434 | D | D | D | D | D |
| PV28435 | D | D | D | D | D |
| PV28436 | D | D | D | D | D |
| PV28440 | D | D | D | D | D |
| PV28441 | D | D | D | D | D |
| PV28437 | D | D | D | D | D |
| PV28438 | D | D | D | D | D |
| PV28444 | D | D | D | D | D |
| PV28443 | D | D | ND | D | D |
| PV28445 | D | D | D | D | D |
| PV28446 | D | D | D | D | D |
| PV28447 | D | D | D | D | D |
| PV28448 | D | D | D | D | D |
| PV28449 | D | D | D | D | D |
| PV28450 | D | D | ND | D | D |
| PV28451 | D | D | ND | D | D |
| PV28452 | D | D | D | D | D |
| PV28453 | D | D | D | D | D |
| PV28454 | D | D | D | D | D |
| PV28455 | D | D | D | D | D |
| PV28459 | D | D | D | D | ND |
| PV28456 | D | D | D | D | D |
| PV28457 | D | D | D | D | D |
| PV28458 | D | D | D | D | D |
| PV28460 | D | D | D | D | D |
| PV28462 | D | D | D | D | D |
| PV28463 | D | D | D | D | D |
| PV28464 | D | D | D | D | D |
| PV28466 | D | D | D | D | D |
| PV28467 | D | D | D | D | D |
| PV28475 | D | D | D | D | D |
| PV28473 | D | D | D | D | D |
| PV28474 | D | D | D | D | D |
| PV28477 | D | D | D | D | D |
| PV28478 | D | D | D | D | ND |
| PV28479 | D | D | D | D | D |
| PV28480 | D | D | D | D | D |
| PV28482 | D | D | D | D | D |
| PV28483 | D | D | D | D | D |
| PV28484 | D | D | D | D | D |
| PV28485 | D | D | D | D | ND |
| PV28487 | D | D | D | D | D |
| PV28490 | D | D | D | D | D |
| PV28489 | D | D | D | D | D |
| PV28492 | D | D | D | D | D |
| PV28497 | D | D | D | D | D |
| PV28495 | D | D | D | D | D |
| PV28496 | D | D | D | D | D |
| PV28501 | D | D | D | D | D |
| PV28502 | D | D | D | D | D |
| PV28503 | D | D | D | D | D |
| PV28499 | D | D | D | D | D |
| PV28500 | D | D | ND | D | D |
| PV28504 | D | D | D | D | D |
| PV28507 | D | D | ND | D | ND |
| PV28506 | D | D | D | D | D |
| PV28508 | D | D | ND | D | D |
| PV29137 | D | D | ND | ND | ND |
| PV29138 | D | D | D | D | D |
| PV29143 | D | D | D | D | D |
| PV29139 | D | D | D | D | D |
| PV29140 | D | D | D | D | ND |
| PV29141 | D | D | D | D | D |
| PV29144 | D | D | D | D | D |
| PV29142 | D | D | D | D | D |
| PV29155 | D | D | D | D | D |
| PV29153 | D | D | D | D | D |
| PV29154 | D | D | D | D | D |
| PV29159 | D | D | D | D | D |
| PV29157 | D | D | D | D | D |
| PV29158 | D | D | ND | D | D |
| PV29160 | D | D | D | D | D |
| PV29169 | D | D | D | D | D |
| PV29170 | D | D | D | D | D |
| PV29171 | D | D | D | D | D |
| PV29172 | D | D | D | D | D |
| PV29174 | D | D | ND | D | D |
| PV29173 | D | ND | ND | D | D |
| PV29175 | D | D | D | D | D |
| PV29176 | D | D | D | D | D |
| PV29185 | D | D | D | D | D |
| PV29188 | D | D | ND | D | D |
| PV29190 | D | D | D | D | D |
| PV29192 | D | D | ND | D | D |
| PV29203 | D | D | D | D | D |
| PV29204 | D | D | D | D | D |
| PV29206 | D | D | D | D | D |
| PV29208 | D | D | D | D | ND |
| PV29219 | D | D | D | D | D |
| PV29222 | D | D | D | D | D |
| PV29220 | D | D | D | D | D |
| PV29223 | D | D | D | D | D |
| PV29221 | D | D | D | D | D |
| PV36897 | D | D | D | D | D |
| PV36898 | ND | D | D | D | D |
| PV36901 | D | D | D | D | D |
| PV36902 | D | D | D | D | D |
| PV36904 | D | ND | D | D | ND |
| PV35332 | D | D | ND | D | ND |
| PV35333 | D | D | D | D | D |
| PV35334 | D | D | D | D | D |
| PV35336 | D | D | D | D | D |
| PV35287 | D | D | D | D | D |
| PV35288 | D | D | D | D | D |
| PV35289 | D | D | D | D | D |
| PV35290 | D | D | D | D | D |
| PV35295 | D | D | D | D | ND |
| PV35292 | D | D | D | D | D |
| PV35293 | D | D | D | D | ND |
| PV35297 | D | D | D | D | D |
| PV35291 | D | D | D | D | D |
| PV35299 | D | D | D | D | D |
| PV35300 | D | D | ND | D | ND |
| PV35298 | D | D | D | D | ND |
| PV35294 | D | D | D | D | D |
| PV35301 | D | D | D | D | ND |
| PV35339 | D | D | D | D | ND |
| PV35337 | D | D | D | D | D |
| PV35338 | D | D | D | D | ND |
| PV35340 | D | D | D | D | ND |
| PV35302 | D | D | D | D | D |
| PV35303 | D | D | D | D | D |
| PV35305 | D | D | D | D | D |
| PV35304 | D | D | D | D | D |
| PV35307 | D | D | D | D | D |
| PV35309 | D | D | D | D | D |
| PV35310 | D | D | D | D | D |
| PV35317 | D | D | D | D | D |
| PV35313 | D | D | D | D | D |
| PV35314 | D | D | D | D | D |
| PV35316 | D | D | D | D | D |
| PV35319 | D | D | ND | D | D |
| PV35323 | D | D | D | D | D |
| PV35321 | D | D | D | D | ND |
| PV35322 | D | D | D | D | D |
| PV35325 | D | D | D | D | D |
| PV35324 | D | ND | D | D | ND |
| PV35326 | D | D | D | D | D |
| PV35345 | D | D | D | D | D |
| PV35341 | D | D | D | D | ND |
| PV35149 | D | D | D | D | ND |
| PV35343 | D | D | D | D | ND |
| PV35327 | D | D | D | D | D |
| PV35344 | D | D | D | D | D |
| PV35347 | D | D | D | D | D |
| PV35328 | D | D | D | D | D |
| PV35346 | D | ND | ND | D | D |
| PV35348 | D | D | D | D | D |
| PV35349 | D | ND | D | D | D |
| PV35350 | D | ND | ND | D | D |
| PV35194 | D | D | D | D | D |
| PV35331 | D | D | D | D | D |
| PV35662 | D | D | D | D | D |
| PV35673 | D | D | ND | D | D |
| PV35666 | D | D | D | D | D |
| PV35667 | D | ND | ND | D | ND |
| PV35668 | D | ND | D | D | D |
| PV35669 | D | D | D | D | D |
| PV35664 | D | D | D | D | D |
| PV35680 | D | D | D | D | D |
| PV35682 | D | D | D | D | D |
| PV35678 | D | D | D | D | D |
| PV35679 | D | D | D | D | D |
| PV35685 | D | D | D | D | D |
| PV35686 | D | D | D | D | D |
| PV35683 | D | D | D | D | D |
| PV35684 | D | D | D | D | D |
| PV35696 | D | D | D | D | D |
| PV35695 | D | D | ND | D | D |
| PV35697 | D | D | D | D | D |
| PV35699 | D | D | D | D | D |
| PV35698 | D | D | D | D | D |
| PV35702 | D | D | D | D | D |
| PV35704 | D | D | ND | D | D |
| PV35706 | D | D | D | D | D |
| PV35707 | D | D | D | D | D |
| PV35705 | D | D | D | D | D |
| PV35709 | D | D | D | D | D |
| PV35714 | D | D | D | D | D |
| PV35713 | D | D | D | D | D |
| PV35715 | D | D | D | D | D |
| PV35716 | D | D | D | D | D |
| PV35717 | D | D | D | D | D |
| PV35719 | D | D | D | D | D |
| PV35720 | D | D | D | D | D |
| PV35722 | D | D | D | D | D |
| PV35917 | D | D | ND | ND | ND |
| PV35918 | D | D | ND | D | ND |
| PV35919 | D | D | D | D | D |
| PV35921 | D | D | ND | D | D |
| PV35923 | D | D | D | D | D |
| PV35924 | D | D | D | D | D |
| PV35925 | D | D | D | D | D |
| PV35926 | D | D | D | D | ND |
| PV35927 | D | D | D | D | ND |
| PV35928 | D | D | ND | D | D |
| PV35929 | D | D | D | D | D |
| PV35930 | D | D | D | D | D |
| PV35931 | ND | D | D | D | D |
| PV35935 | D | D | D | D | ND |
| PV35933 | D | D | D | D | ND |
| PV35934 | D | D | D | D | D |
| PV35939 | D | ND | D | D | ND |
| PV35932 | D | D | ND | D | ND |
| PV35967 | D | D | D | D | D |
| PV35940 | D | D | D | D | D |
| PV35941 | D | D | D | D | D |
| PV35936 | D | D | D | D | ND |
| PV35937 | D | D | D | D | ND |
| PV35942 | D | D | D | D | D |
| PV35938 | D | D | D | D | D |
| PV35943 | D | D | D | D | D |
| PV35944 | D | D | D | D | D |
| PV35945 | D | D | D | D | D |
| PV35947 | D | D | ND | D | D |
| PV35948 | D | D | ND | D | D |
| PV35946 | D | D | D | D | D |
| PV35949 | D | D | D | D | ND |
| PV35953 | D | D | D | D | D |
| PV35954 | D | D | D | D | D |
| PV35955 | D | D | D | D | D |
| PV35957 | D | D | D | D | D |
| PV35962 | D | D | D | D | D |
| PV35963 | D | D | D | D | D |
| PV35958 | D | D | ND | D | D |
| PV35959 | D | D | D | D | D |
| PV35960 | D | D | D | D | D |
| PV35970 | D | D | D | D | D |
| PV35961 | D | D | D | D | D |
| PV35965 | D | D | D | D | D |
| PV35966 | D | D | D | D | D |
| PV35971 | D | D | ND | D | D |
| PV35972 | D | D | D | D | D |
| PV35973 | D | D | ND | ND | ND |
| PV35974 | D | D | ND | D | D |
| PV35975 | D | D | D | D | D |
| PV35977 | D | D | D | D | D |
| PV35976 | D | ND | D | D | D |
| PV35990 | D | D | ND | D | ND |
| PV35980 | D | D | D | D | D |
| PV35981 | D | D | D | D | D |
| PV35982 | ND | D | D | D | D |
| PV35978 | D | D | D | D | D |
| PV35979 | D | D | D | D | D |
| PV35983 | D | D | D | D | D |
| PV35989 | D | D | D | D | D |
| PV35985 | D | D | D | D | D |
| PV35984 | D | D | D | D | D |
| PV35991 | D | D | D | D | D |
| PV35992 | D | D | ND | D | ND |
| PV35993 | D | D | D | D | D |
| PV35995 | D | D | D | D | D |
| PV36003 | D | D | D | D | D |
| PV35996 | D | ND | ND | D | ND |
| PV35997 | D | D | D | D | D |
| PV35998 | D | D | D | D | D |
| PV36023 | D | D | ND | D | D |
| PV36024 | D | D | D | D | D |
| PV36026 | D | D | D | D | D |
| PV36025 | D | D | ND | D | D |
| PV36027 | D | D | D | D | D |
| PV36032 | D | D | D | D | D |
| PV36048 | D | D | ND | D | D |
| PV36036 | D | D | ND | ND | ND |
| PV36037 | D | D | ND | D | D |
| PV36049 | D | D | D | D | ND |
| PV36046 | D | D | ND | D | D |
| PV36038 | D | ND | D | D | D |
| PV36039 | D | ND | D | D | D |
| PV36040 | D | ND | D | D | D |
| PV36042 | D | ND | D | D | D |
| PV36043 | D | ND | D | D | D |
| PV36047 | D | ND | D | D | ND |
| PV36052 | D | D | D | D | D |
| PV36057 | D | D | D | D | D |
| PV36060 | D | D | D | D | D |
| PV36058 | D | D | D | D | D |
| PV36059 | D | D | ND | D | D |
| PV36061 | D | D | ND | D | D |
| PV36056 | D | D | D | D | D |
| PV36062 | D | ND | D | D | D |
| PV36067 | D | D | D | D | D |
| PV36063 | D | D | D | D | D |
| PV36066 | D | D | D | D | D |
| PV36070 | D | D | D | D | D |
| PV36071 | D | D | D | D | D |
| PV36080 | D | ND | D | D | D |
| PV36069 | D | D | D | D | D |
| PV36073 | D | D | ND | D | D |
| PV36081 | D | ND | ND | D | D |
| PV36079 | D | D | D | D | D |
| PV36074 | D | D | D | D | D |
| PV36075 | D | D | D | D | D |
| PV36076 | D | D | D | D | D |
| PV36082 | D | ND | D | D | D |
| PV36077 | D | D | D | D | D |
| PV36084 | D | ND | D | D | D |
| PV36083 | D | D | D | D | D |
| PV36085 | D | D | ND | D | ND |
| PV36086 | D | D | D | D | D |
| PV36087 | D | D | D | D | D |
| PV36088 | D | D | D | D | D |
| PV36089 | D | D | D | D | D |
| PV36090 | D | D | ND | D | ND |
| PV36091 | D | ND | D | D | D |
| PV36093 | D | D | D | D | D |
| PV36094 | D | D | ND | D | D |
| PV36096 | D | D | ND | ND | ND |
| PV36097 | D | D | D | D | D |
| PV36098 | D | D | D | D | D |
| PV36101 | D | D | ND | D | ND |
| PV36103 | D | D | D | D | D |
| PV36102 | D | D | D | D | D |
| PV36104 | D | D | D | D | D |
| PV36105 | D | D | D | D | D |
| PV36107 | D | D | D | D | ND |
| PV36106 | D | ND | ND | D | ND |
| PV36110 | D | D | D | D | D |
| PV36402 | D | D | D | D | D |
| PV36400 | D | D | D | D | D |
| PV36401 | D | D | ND | D | D |
| PV36461 | D | D | ND | D | D |
| PV36462 | D | ND | D | D | ND |
| PV36404 | D | ND | ND | D | D |
| PV36403 | D | D | D | D | D |
| PV36397 | D | D | D | D | D |
| PV36406 | D | D | D | D | D |
| PV36407 | D | D | D | D | D |
| PV36408 | D | D | D | D | ND |
| PV36411 | D | D | D | D | D |
| PV36409 | D | D | D | D | D |
| PV36412 | D | D | D | D | ND |
| PV36410 | D | D | D | D | D |
| PV36414 | D | D | ND | D | D |
| PV36415 | D | D | D | D | D |
| PV36427 | ND | D | D | D | D |
| PV36422 | D | D | D | D | D |
| PV36416 | D | D | D | D | D |
| PV36426 | D | D | D | D | D |
| PV36419 | D | D | D | D | ND |
| PV36420 | D | D | D | D | D |
| PV36428 | D | ND | D | D | ND |
| PV36429 | D | D | D | D | D |
| PV36430 | D | D | D | D | ND |
| PV36431 | D | D | D | D | D |
| PV36434 | D | D | ND | D | D |
| PV36432 | D | D | D | D | D |
| PV36435 | D | D | D | D | D |
| PV36446 | D | D | ND | D | ND |
| PV36436 | D | D | ND | D | ND |
| PV36439 | D | D | D | D | D |
| PV36438 | D | D | D | D | ND |
| PV36440 | D | D | D | D | D |
| PV36447 | D | D | D | D | D |
| PV36448 | D | D | ND | D | D |
| PV36449 | D | D | D | D | ND |
| PV36442 | D | D | D | D | D |
| PV36441 | D | D | D | D | D |
| PV36450 | D | D | D | D | ND |
| PV36454 | D | D | ND | D | D |
| PV36453 | D | ND | D | D | ND |
| PV36455 | D | D | D | D | D |
| PV36456 | D | ND | D | D | D |
| PV36458 | D | D | D | D | ND |
| PV36791 | D | ND | ND | D | ND |
| PV36644 | D | D | D | D | D |
| PV36640 | D | D | D | D | D |
| PV36645 | D | D | D | D | D |
| PV36639 | D | D | D | D | D |
| PV36648 | D | D | ND | D | D |
| PV36646 | D | D | D | D | D |
| PV36641 | D | D | D | D | D |
| PV36642 | D | D | D | D | D |
| PV36647 | D | D | D | D | D |
| PV36649 | D | D | D | D | D |
| PV36652 | D | ND | ND | D | ND |
| PV36653 | D | D | D | D | D |
| PV36654 | ND | D | D | D | D |
| PV36650 | D | D | D | D | ND |
| PV36651 | D | D | D | D | D |
| PV36655 | D | D | D | D | D |
| PV36656 | D | D | D | D | D |
| PV36659 | D | D | D | D | D |
| PV36660 | D | D | D | D | D |
| PV36661 | D | D | D | D | D |
| PV36663 | D | ND | D | D | D |
| PV36664 | D | D | ND | D | D |
| PV36666 | D | D | D | D | D |
| PV36670 | D | D | D | D | ND |
| PV36671 | D | D | D | D | D |
| PV36672 | D | D | D | D | D |
| PV36673 | D | D | D | D | D |
| PV36674 | D | D | D | D | D |
| PV36677 | D | D | D | D | D |
| PV36678 | D | D | ND | D | D |
| PV36689 | D | D | D | D | D |
| PV36679 | D | ND | ND | D | D |
| PV36680 | D | D | ND | D | D |
| PV36681 | D | D | ND | D | D |
| PV36682 | D | D | D | D | D |
| PV36685 | D | D | ND | D | D |
| PV36684 | D | D | D | D | D |
| PV36687 | D | D | ND | D | D |
| PV36683 | D | D | ND | D | ND |
| PV36688 | D | D | ND | D | D |
| PV36690 | D | D | D | D | D |
| PV36691 | D | D | D | D | D |
| PV36692 | D | D | D | D | D |
| PV36693 | D | D | D | D | D |
| PV36694 | D | D | D | D | D |
| PV36889 | D | D | D | D | D |
| PV36890 | D | D | ND | D | D |
| PV36932 | D | D | D | D | D |
| PV36951 | D | D | ND | D | D |
| PV36945 | D | D | D | D | D |
| PV36946 | D | D | ND | D | D |
| PV36947 | D | D | D | D | D |
| PV36948 | D | D | D | D | D |
| PV36949 | D | D | D | D | D |
| PV36950 | D | D | D | D | ND |
| *Detected, D; Not detected, ND | | | | | |

**Supplemental Figure S1. SARS-CoV-2 positional mismatches at target primer/probe binding sites (continued).** Same as Figure 3 for the ORF1AB target.

**Supplemental Figure S2. Substitutions in non-B.1.1.7 genomes associated with N3 target dropout.** Same as Figure 4 for genomes (n = 45) that do not map to B.1.1.7 and have dropout of N3 target detection. Note, substitutions in the N3 forward PBS other than the A28095T substitution are depicted and highlighted in blue with white typeface font.
