## Supplementary figures and images for "Robust clinical detection of SARS-CoV-2 variants by RT-PCR/MALDI-TOF multi-target approach"

### Fig. S1

#### Supplemental Figure S1

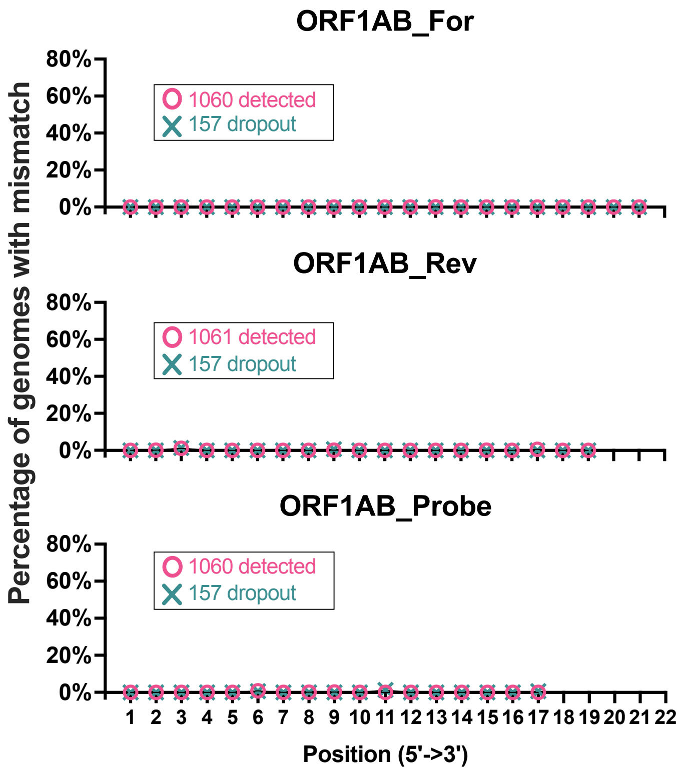

### Fig. S2

#### Supplemental Figure S2

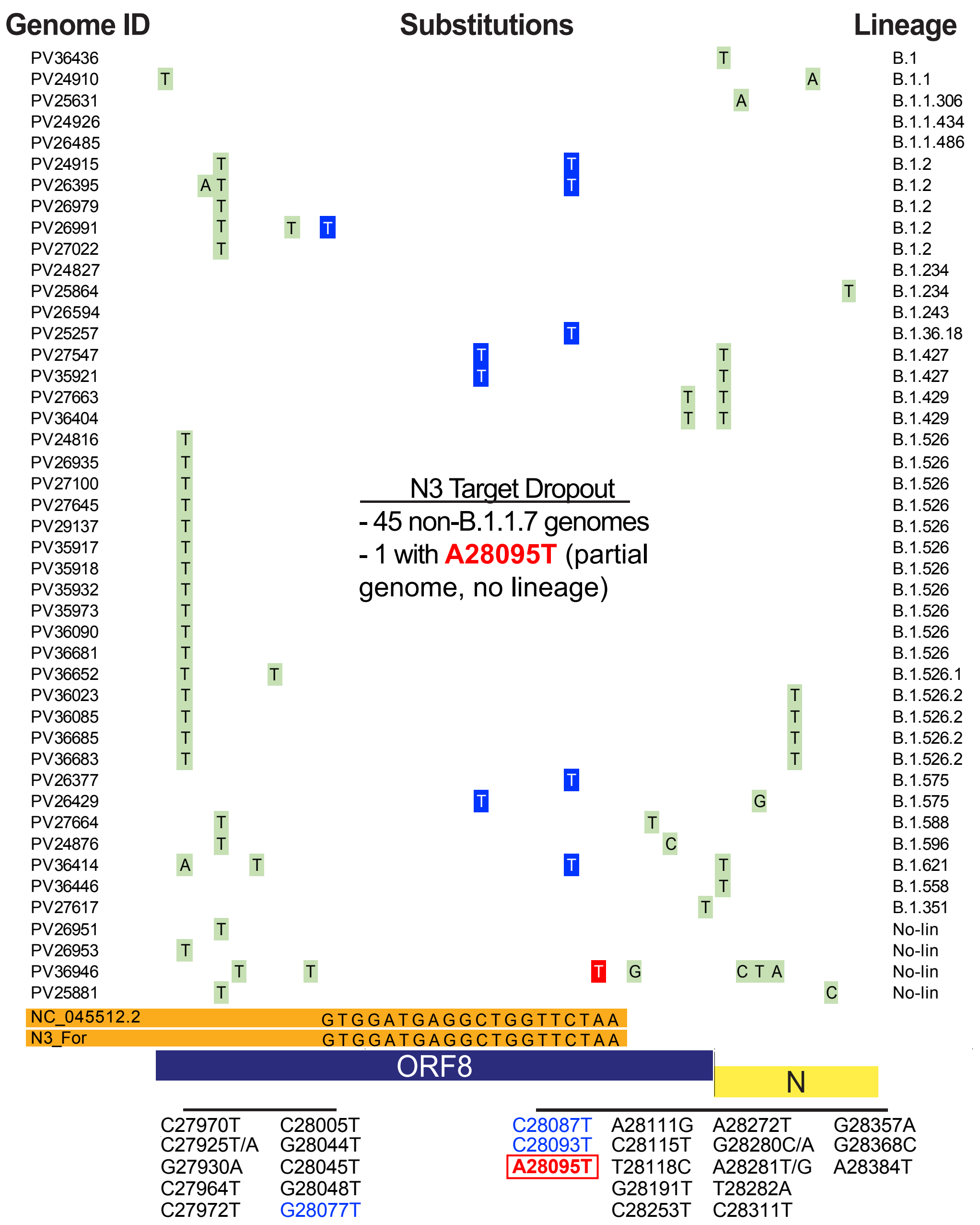
